# *CXCL12* drives natural variation in coronary artery anatomy across diverse populations

**DOI:** 10.1101/2023.10.27.23297507

**Authors:** Pamela E. Rios Coronado, Daniela Zanetti, Jiayan Zhou, Jeffrey A. Naftaly, Pratima Prabala, Azalia M. Martínez Jaimes, Elie N. Farah, Xiaochen Fan, Soumya Kundu, Salil S. Deshpande, Ivy Evergreen, Pik Fang Kho, Austin T. Hilliard, Sarah Abramowitz, Saiju Pyarajan, Daniel Dochtermann, Million Veteran Program, Scott M. Damrauer, Kyong-Mi Chang, Michael G. Levin, Virginia D. Winn, Anca M. Paşca, Mary E. Plomondon, Stephen W. Waldo, Philip S. Tsao, Anshul Kundaje, Neil C. Chi, Shoa L. Clarke, Kristy Red-Horse, Themistocles L. Assimes

**Author notes:** Pamela E. Rios Coronado, Daniela Zanetti, and Jiayan Zhou contributed as co-lead junior authors. Shoa L. Clarke, Kristy Red-Horse, and Themistocles L. Assimes contributed as co-supervising senior authors.

## Abstract

To efficiently distribute blood flow to cardiac muscle, the coronary artery tree must follow a specific branching pattern over the heart. How this pattern arises in humans is unknown due to the limitations of studying human heart development. Here, we leveraged a natural variation of coronary artery anatomy, known as coronary dominance, in genetic association studies to identify the first known driver of human coronary developmental patterning. Coronary dominance refers to whether the right, left, or both coronary arteries branch over the posterior left ventricle, but whether this variability is heritable and how it would be genetically regulated was completely unknown. By conducting the first large-scale, multi-ancestry genome-wide association study (GWAS) of coronary dominance in 61,043 participants of the VA Million Veteran Program, we observed moderate heritability (27.7%) with ten loci reaching genome wide significance. An exceptionally strong association mapped DNA variants to a non-coding region near the chemokine *CXCL12* in both European and African ancestries, which overlapped with variants associated with coronary artery disease. Genomic analyses predicted these variants to impact *CXCL12* levels, and imaging revealed dominance to develop during fetal life coincident with *CXCL12* expression. Reducing *Cxcl12* in mice to model the human genetics altered septal artery dominance patterns and caused coronary branches to develop away from *Cxcl12* expression domains. *Cxcl12* heterozygosity did not compromise overall artery coverage as seen with full deletion, but instead changed artery patterning, reminiscent of the human scenario. Together, our data support *CXCL12* as a critical determinant of human coronary artery growth and patterning and lay a foundation for the utilization of developmental pathways to guide future precision ‘medical revascularization’ therapeutics.

## Introduction

The heart is a muscular organ that beats continuously throughout life. It requires a highly optimized network of coronary arteries to ensure efficient and constant perfusion with oxygenated blood^1,2^. Diseases of the coronary arteries that interrupt blood flow are a leading cause of death worldwide^3^, yet we have little knowledge of how these important vessels are formed and patterned in human hearts.

Most of the major coronary artery branches follow a standard topological pattern in nearly all humans. The left main coronary artery (LCA) originates from the left side of the aorta and gives rise to the left anterior descending (LAD) and left circumflex (LCx) arteries, which provide blood flow to the anterior and lateral walls of the left ventricle, respectively. The right coronary artery (RCA) takes off from the right side of the aorta and follows a course around the right ventricle towards the back of the heart. Except for coronary anomalies, which can cause adverse cardiovascular outcomes, the origins and paths of the LMCA, LAD, LCx, and RCA, are the same throughout the population.

One exception to this standardized anatomy involves the posterior descending artery (PDA), which provides blood to the back of the heart, and variations in structure are termed coronary artery dominance^4^. In approximately 80% of the population, the PDA arises from the RCA (right dominant), but in ∼10%, the RCA is diminutive, and the PDA instead arises from the LCx (left dominant). In the remaining ∼10%, branches perfusing the inferior wall arise from both the RCA and LCx (co-dominant) (**Figure 1A**). An analogous normal variation in artery anatomy has been described in mice involving the origin of the septal artery, which can also be right, left, or co-dominant^5^. Crosses between different strains showed that mouse dominance is genetically regulated^5^, and like humans, right dominance is by far the most common phenotype^5-7^. However, for both humans and mice, how and when dominance is determined is unknown.

**Figure 1.**
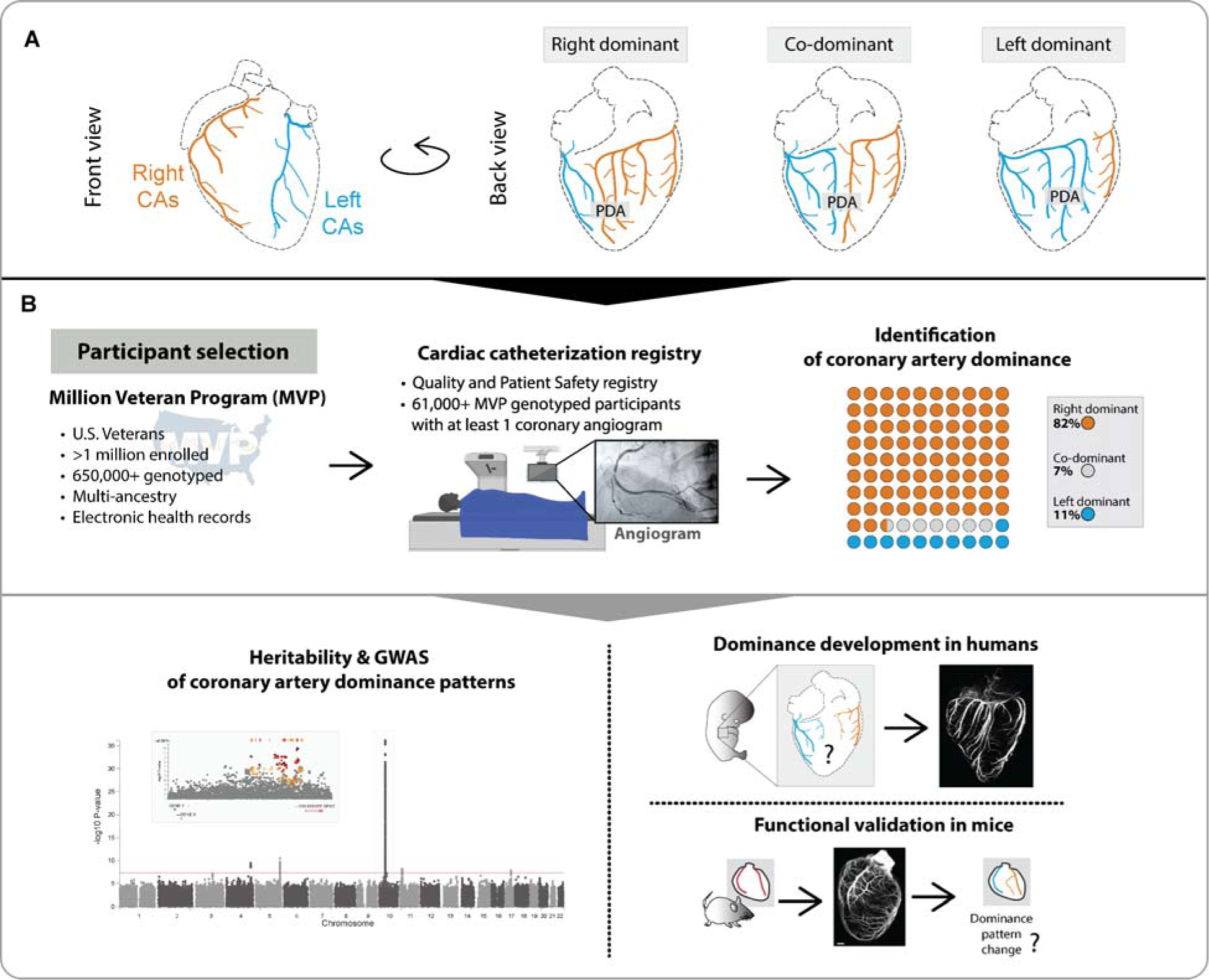
Study design to identify and understand the genetic determinants of coronary artery dominance. (**A**) Variation in the posterior descending artery (PDA) determines dominance in the human heart. (**B**) A multi-ancestry genome-wide association study (GWAS) was performed using genotype data from participants in MVP whose coronary arteries were imaged via angiogram. Analyses were conducted in all participants combined and in cohorts stratified by genetically inferred ancestry. Whole-organ immunolabeling of fetal hearts investigated when dominance is established, and mouse studies validated the most significant genetic association. CAs, coronary arteries.

Developmental biology research has demonstrated that artery development has conserved features across organs and species^8-10^. Work in model organisms has demonstrated that arteries, including coronary arteries, are generated through maturation of an immature capillary plexus. Specifically, endothelial cells (ECs) of the immature capillary plexus undergo artery differentiation, migrate against the direction of blood flow, and coalesce together to progressively lengthen artery branches^11-14^. During this process, the artery ECs induce surrounding pericytes or mesenchyme to differentiate into smooth muscle^15,16^. Thus, human coronary artery development and dominance establishment could involve some of the molecular pathways known to drive these processes in model organisms.

To study the mechanisms of human coronary artery developmental patterning, we took advantage of the natural variation in coronary dominance to conduct the first genome-wide association study (GWAS) of coronary anatomical phenotypes. We analyzed more than 61,000 genotyped participants of the Million Veteran Program (MVP) who had phenotypic data from invasive coronary angiograms. We found significant heritability and identified *CXCL12 -* a gene implicated in coronary artery disease (CAD) over sixteen years ago^17^ - as a potential regulator of coronary patterning. Genomics analyses nominated candidate causal variants in the *CXCL12* locus, and *in vivo* mouse experiments supported a functional role in influencing coronary artery localization and dominance during heart development.

## Results

### A large-scale, genetically diverse genome-wide association study of coronary artery dominance

We leveraged the Million Veteran Program (MVP) linked to the Veterans Health Administration (VA) Clinical Assessment Reporting and Tracking (CART) Program for cardiac catheterization laboratories to conduct a large-scale, multi-ancestry GWAS of dominance (**Figure 1B**). Within this biobank, we identified 61,043 participants with a dominance assessment and genotype data available imputed to a multi-ancestry TOPMed reference panel (**Figure 1B, Supplementary Table 1**). We used Scalable and Accurate Implementation of GEneralized mixed model (SAIGE)^18^ to conduct each GWAS. Ancestry-specific GWAS were conducted in the three largest genetically inferred ancestry groups: European (EUR), African (AFR), and Admixed American (AMR). We also performed pooled analyses containing all participants (All ancestry), including those not assigned to the major ancestry groups. Because the genetics of dominance had never been explored, it was not known a priori whether left- and co-dominance were related and should be combined or not. Thus, to be thorough and unbiased, we ran two GWAS for each population: (1) right (reference) versus left plus co-dominance (RvsLCo) and (2) right (reference) versus left only (RvsL).

Routine statistical analyses indicated a robust genetic signature underlying coronary dominance. First, quantile-quantile (Q-Q) plots comparing P-values for genetic variants associated with dominance to those expected for no association (null hypothesis) demonstrated a strong genetic signal in the All ancestry (**Figure 2A**), EUR (**Figure 2B**), and AFR (**Figure 2C**) GWAS. There was no evidence of population stratification based on the calculated genomic control lambdas for common variants with minor allele frequency (MAF) > 0.01. These observations were bolstered by a point estimate of total SNP-based narrow-sense heritability (*h*^2^) of 0.277 (95% confidence intervals, 0.150-0.404) among EUR using a linkage disequilibrium (LD) and MAF stratified multicomponent approach implemented in GCTA, GREML-LDMS-I, and assuming the observed prevalence of non-right dominance of 17% (**Supplementary Figure 1A-B**, and **Supplementary Table 2**). A large fraction of the additive genetic variance is derived from low frequency and rare variants (**Supplementary Figure 1C**). The genetic signal was weak to non-existent among AMR (**Figure 2D**), likely due to the lower numbers in this group (**Supplementary Table 1**). Thus, a clear genetic signal emerged for coronary artery dominance in datasets with greater than 2000 non-right individuals.

**Figure 2.**
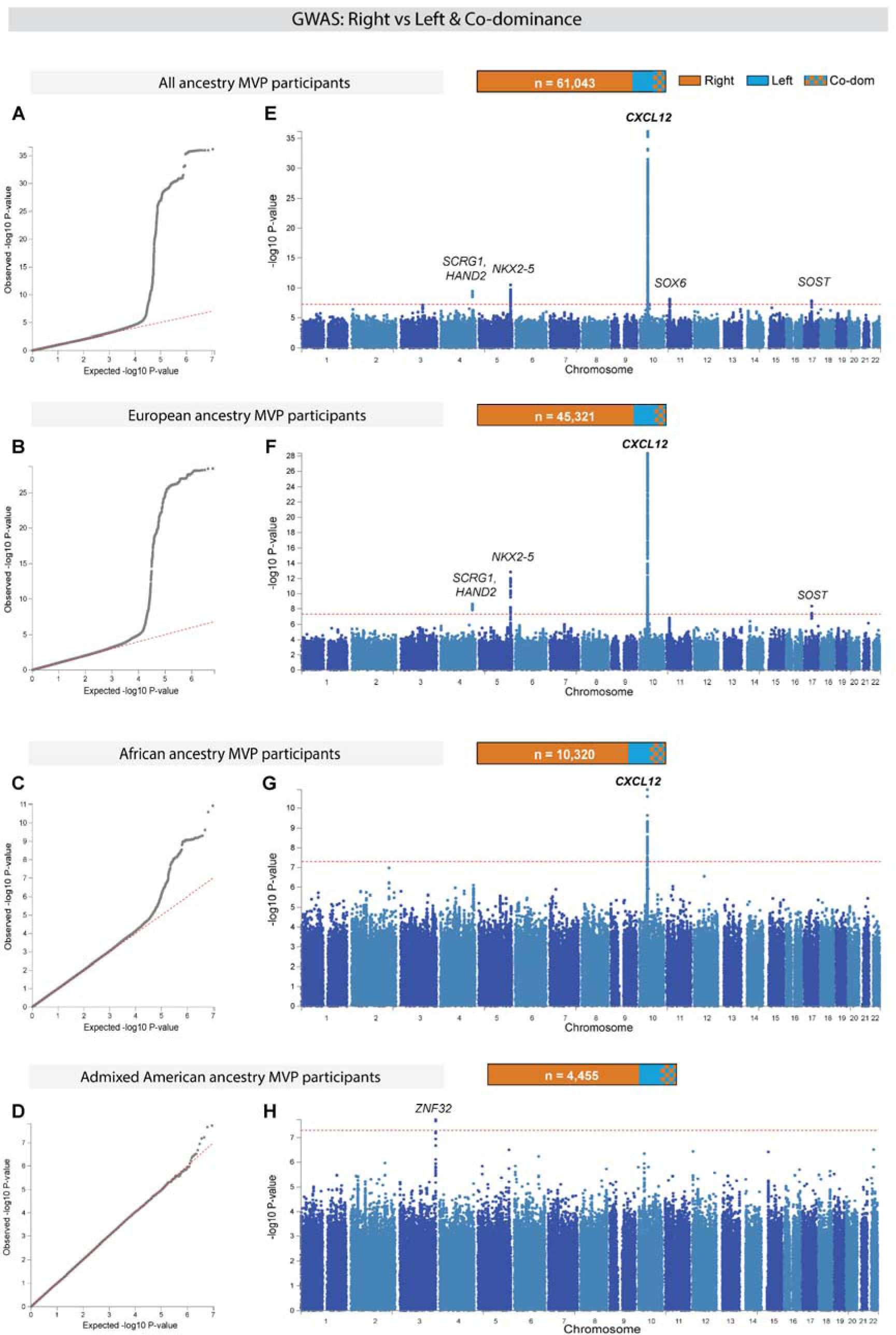
Genome-wide association study (GWAS) of right versus left/co-dominance in the Million Veteran Program (MVP). GWAS was conducted in all participants independent of ancestry (All ancestry) as well as in each of the three largest genetically inferred ancestry subgroups: European, African, and Admixed American. (**A-D**) Quantile-quantile plots (Q-Q) show the observed versus expected P-values from association for all genetic variants. Red line indicates null hypothesis. (**E-H**) Manhattan plots for the corresponding Q-Q plots with P-values of genetic variants. Gene(s) mapped to regions reaching genome wide significance (GWS) are labeled. Red line indicates GWS threshold (P-value<5x10^-8^).

Ten loci across seven chromosomes reached genome wide significance (GWS) in one or more of the GWAS, as defined by an algorithm implemented in FUMA-GWAS^19^ (**Figure 2E-H**, **Table 1, Supplementary Figure 2**, and **Supplementary Table 3**). Of these, we consider seven loci of highest confidence because they contained common lead variants in both the All ancestry GWAS and in one or more of the ancestry specific GWAS (**Table 1**). Two additional loci were defined by either one or a small number of rare variants in high LD with MAF < 0.005. One locus on chromosome 3 was defined by multiple low frequency variants reaching GWS only in AMR despite higher frequencies and better power to implicate the same region in the All ancestry, EUR, and AFR groups. Furthermore, the saddle point approximation (SPA) for P-values of SNPs found in this region for AMR did not converge in the other populations.

**Table 1.**
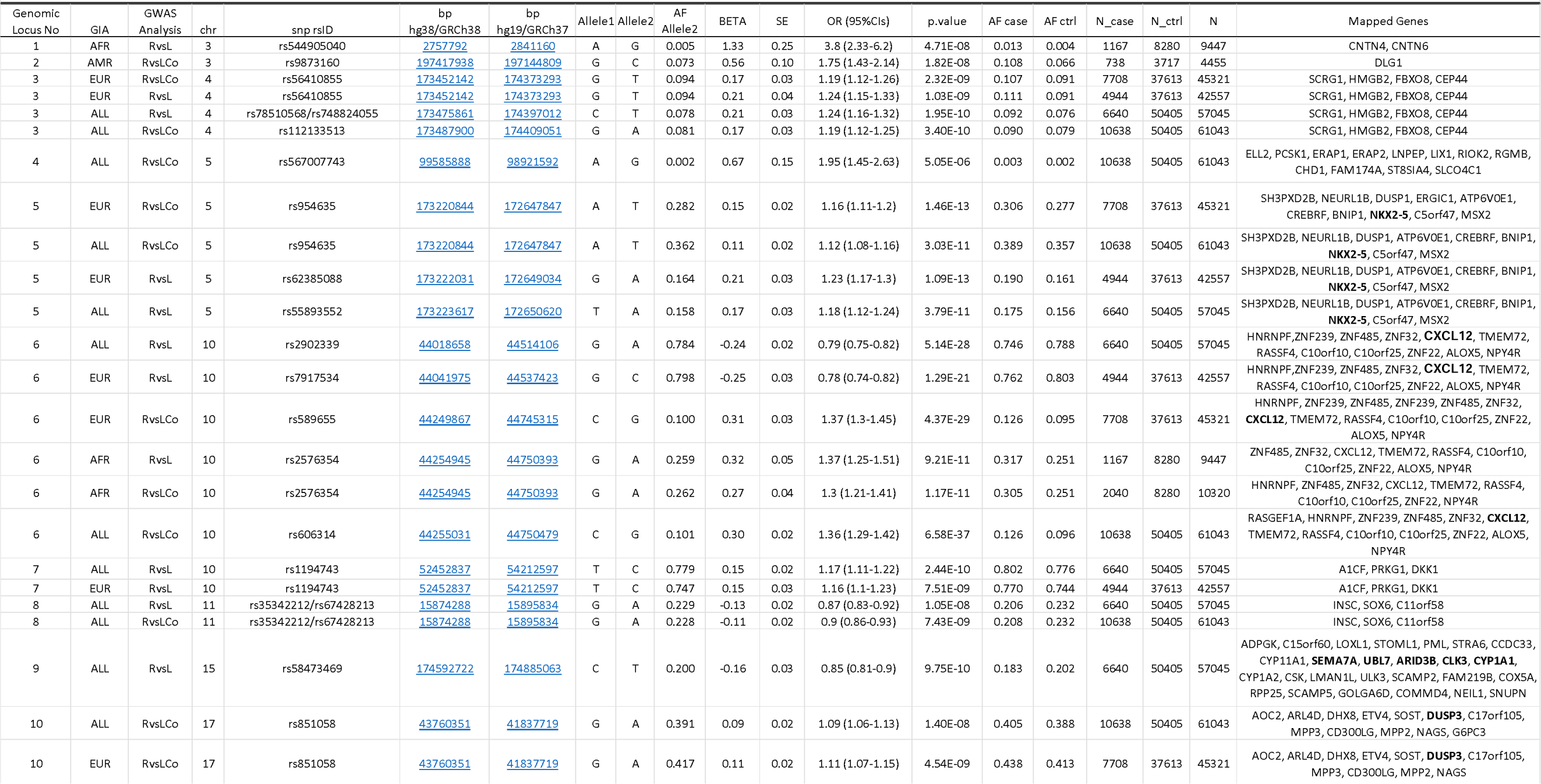
Lead genetic variants, effect sizes, allele frequencies, and mapped genes at loci reach genome wide significance for coronary artery dominance status. Genomic locus No: Index of genomic risk loci, GIA: Genetically inferred ancestry (ALL; all ancestral groups combined, EUR; genetically inferred European subjects, AFR; genetically inferred African subjects, AMR; genetically inferred Admixed-Americans), GWAS: Genome-wide assocociation analysis comparison (RvsLCo; right dominance as reference versus left and co-dominant subjects combined, RvsL; right dominance (reference) versus left dominance subjects excluding subjects with mixed dominance, chr: chromosome, SNP rsID: variant ID, bp hg38/GRCh38: base position on human build 38, bp hg19/GRCh37: base position on human build 37, Allele1: allele 1, Allele2: allele 2, AF_Allele2: allele frequency of allele 2, BETA: effect size of allele 2, SE: standard error of BETA, OR (95%CIs): Odds ratio and 95% confidence intervals of non-right dominance to right dominance for allele 2 compared to allele1, p.value: p value with SaddlePoint Approximation applied, AF_case: allele frequency of allele 2 in cases, AF_ctrl: allele frequency of allele 2 in controls, N_case: sample size in cases, N_ctrl: sample size in controls, N: sample size cases and controls combined. Mapped genes include either by position (ANNOVAR), gene expression (eQTL), or chromatin interaction. Genes mapped by all three methods are bolded.

Variants were near genes known to play a role in cardiac development (e.g., *NKX2-5*) and the most significant with the highest effect size was downstream of *CXCL12*, which, remarkably, reached GWS in both EUR and AFR (**Figure 2E, F,** and **G**). Regional association and LD plots for GWS loci are included in **Supplementary Figure 3**. Statistical fine mapping with SuSiEx^20^, which is used to help predict potential phenotype-causing variants, identified a single credible set of SNPs in five of the loci, two credible sets within the *NKX2-5* locus on chromosome 5 and four credible sets within the *CXCL12* locus on chromosome 10. **Supplementary Table 4** lists the SNPs within each credible set with the highest posterior probability (PP) for being independently causal. **Supplementary Table 5** provides full SAIGE output for all lead and independently significant variants identified by FUMA, and all SuSiEx lead genetic variants. **Supplementary Table 6** is a list of mapped genes using GTEx v8 and chromatin folding maps as implemented in FUMA. Lastly, **Supplementary Table 7** provides annotation hyperlinks for the lead SNPs in Table 1 to five online variant-based portals. In total, these data identified several loci that merit further examination regarding their involvement in coronary artery patterning.

### Lead genetic variants in both EUR and AFR populations are linked to CXCL12 expression

We next further assessed the *CXCL12* locus because it was the strongest signal observed in multiple GWAS (**Figure 2F** and **G**) and previous studies have demonstrated a role for *Cxcl12* during artery development in model organisms^11,21-25^.

A detailed analysis of the *CXCL12* locus revealed three independent genomic regions containing highly significant variants in the EUR cohort, with two of those being significant in the AFR cohort (**Figure 3A_i_** and **Supplementary Figure 4A, B**). A subgroup analysis of the locus restricted to a comparison of co-dominant individuals alone to right dominant individuals revealed a genome wide significant signal that was nearly identical in shape to the signal observed in the right versus left and the right versus left and co-dominant analyses (**Supplementary Figure 3**). To further validate this locus, we performed genetic association testing with coronary dominance in a small multi-ancestry cohort of 3,141 (75.1% EUR, 22.5% AFR, 2.4% Other) from the Penn Medicine Biobank (PMBB). In PMBB, the strongest common variant association with left/co-dominance at the *CXCL12* locus was at rs559580 in the significant genomic region closest to *CXCL12* (MAF: 0.18, OR: 1.46, p=6.7x10^-5^) (**Figure 3A_ii_)**. Directionality was consistent with results for this variant in the MVP All ancestry GWAS (MAF 0.16, OR: 1.25, p=1.3x10^-28^).

**Figure 3.**
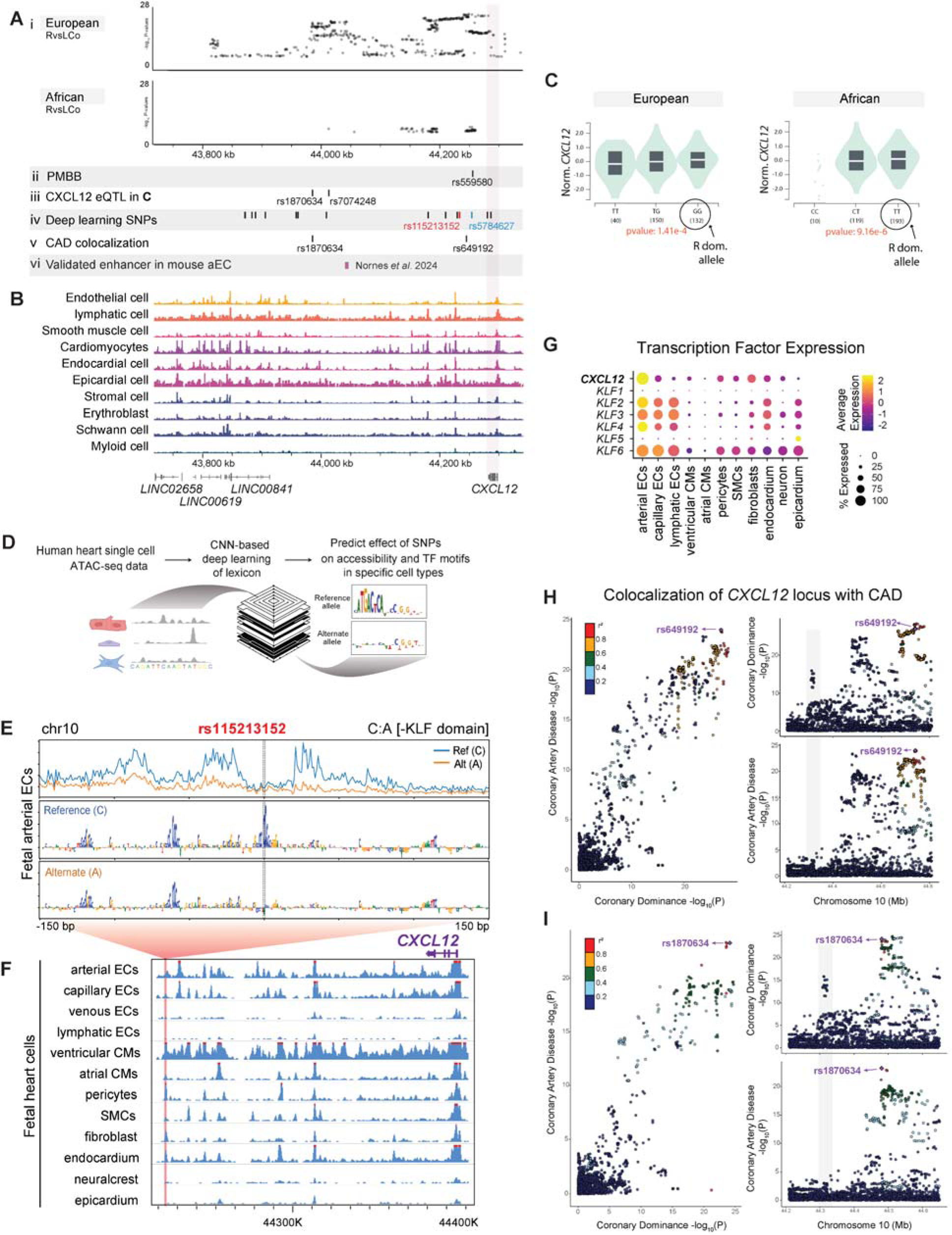
Dominance variants localize to putative *CXCL12* regulatory regions. (**A**) SNPs reaching genome wide significance in the indicated MVP populations aligned with variants from additional analyses: Penn Medicine Biobank (PMBB) replication study, Genotype-Tissue Expression (GTEx) eQTLs, deep learning-based predictions, and coronary artery disease (CAD) colocalization. An *in vivo* validated *Cxcl12* enhancer is also shown. (**B**) Location of accessible chromatin in human fetal heart cell types in relation to dominance SNPs in **A**. (**C**) Violin plots of two lead SNP eQTLs revealed that a higher prevalence of right dominance is associated with increased *CXCL12* in GTEx data. Left is rs1870634; right is rs7074248. Testis expression levels by genotype are shown, but directionality is the same in arterial and coronary tissues (see **Supplementary Figure 5A, B**). (**D**) Schematic of convolutional neural network (CNN) deep learning models used to predict the effect of alternate alleles. (**E**) Predicted per-base accessibility for reference and alternate alleles of rs115213152 suggest a disrupted KLF motif. (**F**) Sequencing tracks of chromatin accessibility near *CXCL12*. Red highlights rs115213152 region where open chromatin coincides with *CXCL12* promoter accessibility in select cardiac cell types, including coronary artery cells. (**G**) ScRNAseq from fetal hearts revealed co-expression of *CXCL12* and *KLFs*, supporting a potential role for these transcription factors in the activity predicted in **E**. (**H** and **I**) LocusCompare ouput displaying colocalization plot (left), and LocusZoom plots for each trait (right). The SNP with the highest H4 posterior probability using COLOC is labelled in purple with additional variants in linkage disequilibrium with labelled SNP colored according to R^2^ value. **Top panel (H)** is unmodified analysis and **bottom panel (I)** shows result when the first colocalization signal at rs649192 (44650000-44850000) is masked.

Significant variants within MVP were intergenic and overlapped with open chromatin regions in fetal heart cell types in which the CXCL12 promoter was also accessible (**Figure 3B**). Some of the variants also overlapped with a recently reported Cxcl12 enhancer that activates expression in mouse and zebrafish arterial endothelial cells (**Figure 3A_vi_**)^26^ We next performed CADD scoring to prioritize the non-coding variants and found multiple variants within each independent genomic region in both EUR and AFR with a score >10 consistent with a high likelihood of the variants being deleterious (**Supplementary Figure 4A** and B)^27,28^. We next identified variants associated with the expression of nearby genes (expression quantitative trait loci, eQTLs). In the EUR GWAS, many variants were eQTLs for *CXCL12* across multiple tissues including arteries, although a few were eQTLs for other genes nearby (**Supplementary Figure 4A** and **Supplementary Figure 5A**). In the AFR GWAS, variants were exclusively eQTLs for *CXCL12* and further identified coronary arteries as a relevant tissue (**Supplementary Figure 4B** and **Supplementary Figure 5B**). Databases utilizing HiC data revealed that dominance-associated eQTLs interacted with *CXCL12* and other nearby promoters among EUR, but only with the CXCL12 promoter in AFR (**Supplementary Figure 4C**). To investigate whether higher or lower expression might be correlated with right or left dominance, we chose eQTLs associated with arterial *CXCL12* expression and observed levels detected with the different alleles. This dataset suggested that the right dominance allele was associated with more *CXCL12* expression (**Figure 3A_iii_** and **C**). Thus, at this locus, *CXCL12* is the strongest candidate for a causal gene influencing coronary artery patterning, and the variants identified here likely function through regulating gene expression.

### Deep learning models predict transcription factor motifs affected by dominance SNPs

The above fine mapping tools are widely used to nominate candidate causal DNA variants associated with GWAS traits, but they primarily incorporate data from whole tissues and lack the cell type specific information required to glean detailed mechanistic information. Deep learning models such as convolutional neural networks (CNNs) trained to predict chromatin accessibility profiles from single cell ATAC-seq data are emerging as powerful methods for predicting allelic effects of GWAS variants^29-33^. When interpreted through the lens of the model, various functional predictions can be made such as the disruption of transcription factor motifs in individual cell types. The models can therefore provide specific hypotheses about a variant’s predicted effect on transcription factor binding and chromatin accessibility and nominate putative cell types of action (**Figure 3D**). Indeed, a base-resolution CNN model trained on human fetal heart scATAC-seq data revealed an enrichment of predicted high effect size de-novo variants from congenital heart disease patients specifically in accessible chromatin in artery endothelial cells and identified key transcription factor binding sites disrupted by disease alleles^33^.

We used these cell-type resolved fetal heart models along with similar models trained on chromatin accessibility profiles from cell types derived from adult hearts to score and identify dominance-associated variants predicted to affect chromatin accessibility in cardiac and vascular cell types. The highest-ranking variant at the *CXCL12* dominance locus, rs115213152, was predicted to disrupt motifs belonging to C2H2 domain transcription factors, particularly those in the KLF family, which play a central role in endothelial cell responses to blood flow^34^ (**Figure 3E**). This variant overlapped with the dominance peak nearest gene *CXCL12* (**Figure 3A_iv_**). This SNP was predicted to affect the accessibility of a regulatory element in various fetal cardiac cell types (**Figure 3F**). Interestingly, these cell types - cardiomyocytes, pericytes, fibroblasts, smooth muscle cells, vascular endothelial cells, and endocardial cells - also exhibit robust CXCL12 promoter accessibility (**Figure 3F**). This correlation implies that *CXCL12* could be the targeted gene of these regulatory elements in the developing and adult heart.

To determine the potential transcription factor influencing this element, we scrutinized the expression of all candidate motif-binding factors, such as members of the KLF and SP families, in a publicly available fetal heart scRNAseq dataset^35^. *KLF2, KLF3, KLF4,* and *KLF6* showed both high expression and cell type specificity aligning closely with that of *CXCL12* (**Figure 3G**). Another high-ranking variant, rs5784627, was predicted to disrupt motifs belonging to bZIP domain transcription factors such as JUN and FOS (**Figure 3A_iv_** and **Supplementary Figure 5C** and D), which were also expressed in cardiac cell types coincident with *CXCL12,* although with less cell type specificity (**Supplementary Figure 5E**). Taken together, these data further implicate dominance-associated variants as regulators of *CXCL12* expression in vascular cells of the heart and suggest potentially relevant transcription factors.

### Colocalization of dominance *CXCL12* variants with coronary artery disease variants

The *CXCL12* region identified in our dominance GWAS highly overlaps with a widely replicated signal for CAD^17^. We used the summary statistics from the Cardiogram*plus*C4D Consortium GWAS^36^ and our GWAS of coronary dominance, analyzing the genomic window where the *CXCL12* locus is located (from 44210000 bp to 44810000 bp GRCh37) to perform colocalization analyses^37^. Colocalization predicted shared variants within the *CXCL12* locus for coronary dominance and CAD (**Figure 3A_v_, H,** and **I**). When considering the full interval of the signal, COLOC calculated a global PP Bayesian factor H4 = 0.855. The SNP with the highest H4 PP was rs649192 but nine other highly correlated SNPs (r2>0.8) had similar H4 probabilities (**Supplementary Table 8A**).

The overlap of the signals for the two phenotypes suggested a possible second colocalization in the more proximal middle peak (**Figure 3H**). We thus masked the distal peak closest to *CXCL12* before running COLOC again and observed rs1870634 as a second shared variant within the middle peak. The SNPs highly correlated to rs1870634 also colocalized (**Figure 3I** and **Supplementary Table 8B**). Thus, there are potentially two shared genetic loci that contribute to both dominance status at birth and CAD development later in life.

In contrast, one of the three peaks furthest away from *CXCL12* associated with dominance in the All ancestry and EUR populations did not colocalize with known CAD loci (**Figure 3H** and **I**, grey shading). Interestingly, this peak does overlap open chromatin regions in developing heart cell types (**Figure 3A_i_** and **B**). Thus, coronary artery development and the establishment of dominance may involve a development-specific regulator of *CXCL12* in addition to those also involved in CAD.

### Coronary dominance is apparent at fetal stages when *CXCL12* is expressed

When dominance is determined is unknown; we hypothesized it would be established during heart development with the formation of coronary arteries. Since angiograms and non-invasive imaging procedures are not performed during healthy pregnancies, we investigated whether dominance is present during fetal stages by performing whole-organ immunofluorescence that allowed visualization of the entire artery tree in 3D (**Figure 4A** and **B**)^38,39^. Tracing primary and secondary branches of the right and left coronary arteries of each heart revealed a pattern reminiscent of the adult anatomy. Left and right primary branches stemmed from the aorta and took a course typical of the LAD, LCx, and RCA in all hearts (**Supplementary Figure 6A-C**).

**Figure 4.**
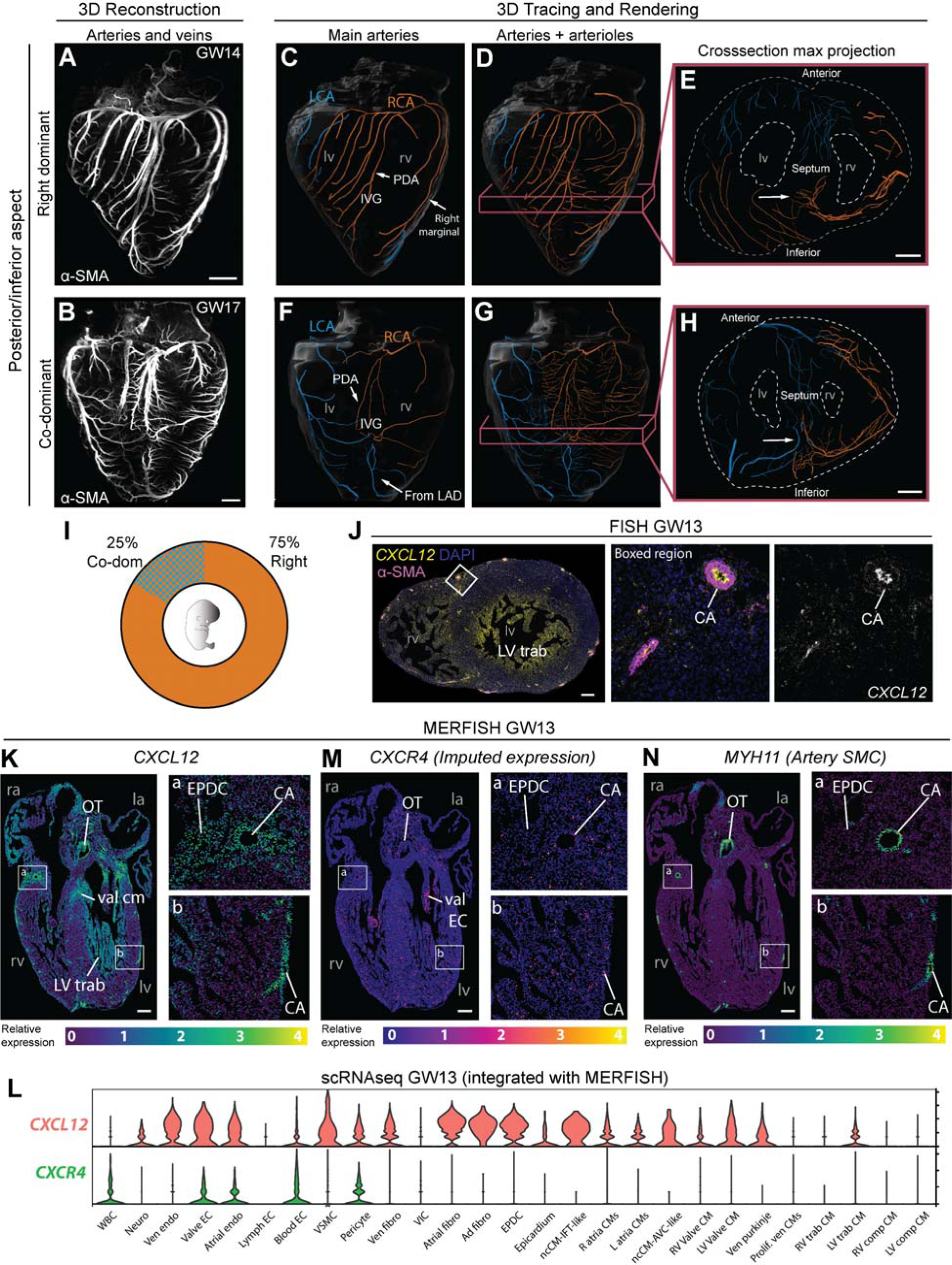
Coronary dominance is apparent during fetal development when *CXCL12* is expressed. (**A** and **B**) Whole-organ immunolabeling of coronary artery smooth muscle (α-SMA) in hearts from the indicated gestational weeks (GW). (**C-H**) Representative artery tracings highlighted branches of the main arteries (**C, F**) or main and lower order arteries (**D, E, G, H**) that originated from the right (RCA, orange) or left (LCA, blue) coronary ostia. (**C-E**) Right dominance was indicated when the artery in the interventricular groove (IVG), i.e., the posterior descending artery (PDA), originated from the RCA (**C** and **D**) and when the inferior septum was occupied by right branches (arrow in **E**). (**F-H**) Co-dominance was indicated when arteries within the IVG stemmed from both the RCA and LCA (**F** and **G**) and the inferior septum was occupied by RCA and LCA branches (arrow in **H**). (**I**) Distribution in a cohort of n=8 human fetal hearts. (**J**) *CXCL12* fluorescence *in situ* hybridization (FISH) and α-SMA immunolabeling on a transverse section through a GW13 heart. Expression was in coronary arteries (CA) and trabecular cardiomyocytes (LV trab). DAPI labeled nuclei. (**K-N**) Spatial expression plots (**K, M, L**) and scRNAseq-derived violin plots (**L**) in GW13 human fetal hearts. MERFISH localized *CXCL12* and *MYH11* while *CXCR4* was imputed by integrating both datasets. ad, adventitial; AVC, atrioventricular; comp, compact; EC, endothelial cell; CM, cardiomyocyte; EPDC, epicardial-derived cells; Endo, endocardial cell; Fibro, fibroblast; IFT, inflow tract; Neuro, neuronal; la, left atrium; Lymph, lymphatic; lv, left ventricle; LV trab, left ventricle trabeculae; ncCM, non-chambered cardiomyocyte; OT, outflow tract; ra, right atrium; RV, right ventricle; trab, trabecular; val, valve; ven, ventricular; VIC, valve interstitial cell; VSMC, vascular smooth muscle; WBC, white blood cell. Scale bars: A, B, E, and J, 500μm; H, 1mm; J, 200μm; K-O, 250μm.

As in adults, there was variability in the extent to which the right and left branches covered the posterior and inferior aspects of the ventricles at gestational weeks (GW) 14-22. Six of the eight hearts (75%) were clearly right dominant in that the interventricular groove had a branch from the right where the adult PDA would be located, and additional right branches extended onto the inferior left ventricle (**Figure 4C**). We also traced a number of lower order arteries and arterioles (**Figure 4D**) and found that the septum was perfused by branches from the right coronary on the inferior side and the LAD on the anterior side (**Figure 4E**). Two of eight hearts (25%) had a different coverage that we scored as co-dominant because both right and left arteries branched into the inferior wall and septum (**Figure 4F-H** and **Supplementary Figure 6D-F**). For example, one heart showed an artery branching onto the interventricular groove from the right, but it only extended halfway down while the rest of the inferior aspect contained branches from the LCx and LAD (**Figure 4F** and **G**). This resulted in the inferior septum being perfused by both right and left (**Figure 4H**). The observation that dominance can be observed as early as GW14 at ratios reminiscent of those in adults (**Figure 4I**), supports a model where it is established during fetal development.

If dominance is established during fetal development and impacted by *CXCL12*, one would predict the expression of this chemokine and its receptor to be near developing coronary arteries during this time. Fluorescence *in situ* hybridization on fetal heart ventricles from GWs 13, 18, and 20 showed expression in coronary arteries, and, at the earliest stage, trabecular cardiomyocytes of the left ventricle (**Figure 4J** and **Supplementary Figure 6G**). Previously generated multiplexed error-robust fluorescence *in situ* hybridization (MERFISH)^40^ data that interrogated the expression of 238 genes across all chambers of a GW13 heart^41^ localized *CXCL12* to multiple regions and cell types. These included coronary arteries, outflow tract smooth muscle, left ventricle trabecular cardiomyocytes, valvular cardiomyocytes, epicardial-derived cells in the coronary sulcus, and others (**Figure 4K**). Single cell RNA sequencing (scRNAseq) from an additional heart at GW13 was integrated with the MERFISH data and showed expression in similar cell types (**Figure 4L**). The integrated datasets were also used to impute expression of *CXCR4,* the gene encoding the CXCL12 receptor. *CXCR4-*positive cells included those nearby *CXCL12* expression, for example, coronary artery endothelial cells and valve endocardial cells near valvular cardiomyocytes (**Figure 4M**). Blood cells and vascular mural cells (pericytes and smooth muscle) also had expression (**Figure 4L**). Coronary artery distribution in these sections is shown with *MYH11*, which marks artery smooth muscle cells (**Figure 4N**). Thus, *CXCL12* is expressed at the right time and place to influence human coronary artery dominance.

### *CXCL12* levels regulate septal artery dominance in mice

To provide functional evidence that changes in *CXCL12* levels could impact coronary branch variation, we analyzed dominance in mice heterozygous for a *Cxcl12* deletion allele. Heterozygous rather than full knockout mice were used to model the decreased expression predicted for human dominance variants, which would not be loss of function. Like humans, mouse coronary arteries have a branch where there is variability across the population. This variation is seen with the mouse-specific SpA, which like the human PDA, can originate from either the right, left, or both coronary arteries^6,7^. In rare instances, the SpA arises directly from the aorta, but still varies in whether it arises from the right, left, or both sides. SpA dominance in mice shares a key feature with human dominance in that the vast majority are right dominant^6,7^. Given these features, and because the cellular mechanisms of artery development are conserved across different organs and species^8^, we considered SpA dominance a good model for studying regulators of human coronary variability.

We assessed SpA dominance in our experiments using either whole-organ immunofluorescence or *in vivo* labeling with fluorescently tagged Isolectin GS-IB_4_ (**Figure 5A** and **Supplementary Figure 7A-C**). Our experiments recapitulated published SpA dominance ratios in both neonatal and adult hearts (**Figure 5B-D** and **Supplementary Figure 7D**), supporting the notion that, like humans, dominance is established early in life^6,7^. For *Cxcl12* haploinsufficiency, the *Cxcl12^DsRed^* mouse strain was utilized. This strain is maintained on the C57Bl/6J background and has part of the *Cxcl12* coding region replaced with a red fluorescent protein, *DsRed*, so that it is both a deletion line and a gene expression reporter^42^. QPCR assessing *Cxcl12* mRNA levels in hearts from wildtype (*Cxcl12^+/+^*), heterozygous (*Cxcl12^DsRed/+^*), and homozygous (*Cxcl12^DsRed/DsRed^*) mice demonstrated that expression in *Cxcl12^DsRed/+^*is approximately 50% lower than *Cxcl12^+/+^* (**Figure 5E**), making them appropriate for assessing SpA dominance in hearts with decreased *Cxcl12* expression.

**Figure 5.**
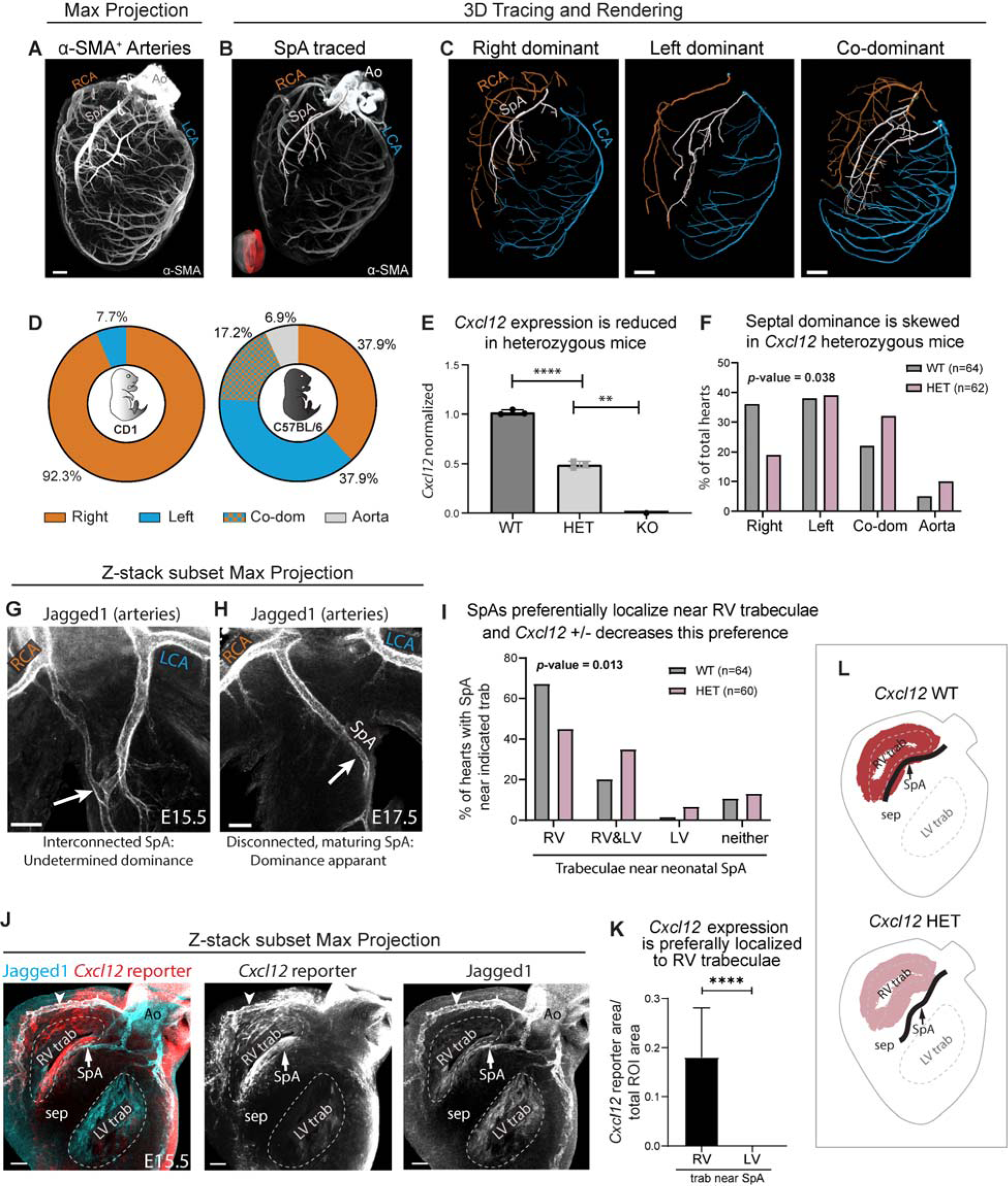
*Cxcl12* influences septal artery dominance in mice. (**A**) Whole-organ immunolabeling of coronary artery smooth muscle (α-SMA) in a postnatal day 6 mouse heart. (**B**) Septal artery (SpA) tracing overlayed onto a Z-stack subset max projection of internal coronary arteries (red region in insert). (**C**) Artery tracings in whole-organ images demonstrating various SpA connections. (**D**) SpA dominance ratios are strain specific. N=13 CD1 neonatal hearts and N=29 for C56BL/6J. (**E**) QPCR using whole hearts from embryonic day (E) 16.5. WT, *Cxcl12^+/+^*; HET, *Cxcl12^DsRed/+^*; KO, *Cxcl12^DsRed/DsRed^*. Each dot is the average value for genotypes within single litters normalized to WT littermates. Error bars, mean±st dev: **, p=0.0027, p=0.0016, p=0.0011; ****, p<0.0001 by two-sided Student’s t-test. (**F**) Blinded scoring of septal artery dominance in WT and HET neonatal mice. P-value by two-tailed Chi-square test. (**G** and **H**) Whole-organ immunolabeling of WT embryonic hearts. First, an immature artery network is attached to both sides (**G**) followed by maturation into a SpA with dominance shortly before birth (**H**). (**I**) Blinded scoring of SpA location as related to trabeculae (see **Supplementary Figure 8I**). P-value by two-tailed Chi-square test. (**J**) *Cxcl12* reporter allele shows that SpA develops in regions of high expression. Arrowhead, expression in artery endothelial cells. (**K**) Reporter allele fluorescence in standardized regions of interest (ROI) along right or left trabeculae. N=14 hearts. Error bars, mean±st dev: ****, p<0.0001 by two-sided Student’s t-test. (**L**) Schematic of *Cxcl12* bias in trabeculae, and HET’s effect on SpA localization. Ao, aorta; LCA, left coronary artery; LV, left ventricle; RCA, right coronary artery; RV, right ventricle; sep, septum; trab, trabecular cardiomyocytes. Scale bars: A, C 400μm; G - I, 100μm.

*Cxcl12^DsRed/+^* mice had shifted SpA dominance ratios when compared with wild type littermates (**Figure 5F**). These heterozygous mice had a lower incidence of right dominance and a higher incidence of co-dominance and aortic-origin. Therefore, *Cxcl12* heterozygosity changed mouse SpA dominance patterns, supporting the hypothesis that genetic variants affecting *CXCL12* expression could influence coronary artery developmental patterning and dominance in humans.

To understand how *Cxcl12* levels influence SpA branching, we first investigated the developmental stages leading to SpA development and dominance establishment and then determined where *Cxcl12* was expressed during this time. Immunolabeling embryonic hearts with the artery endothelial cell marker Jagged1 revealed that Jagged+ vessels appeared in the septum around embryonic days (E)14.5 and 15.5 as a network of interconnected, immature artery precursors that were attached to both the right and left main branches (**Figure 5G** and **Supplementary Figure 8A-C**)^43^. (Note that Jagged1 also sparsely labeled trabecular cardiomyocytes.) This immature artery network is a typical early stage in artery development. Over the next two days, remodeling resulted in progressive development of a major artery within the septum **(Figure 5H).** Notably, most SpAs formed close to the right ventricular trabecular cardiomyocytes that line the chamber lumen—79% grew only alongside the right trabeculae and 18% alongside both while very few (3%) grew only near left trabeculae (**Supplementary Figure 8D, E,** and **F**). This pattern was also observed in neonatal WT hearts (**Figure 5I** and **Supplementary Figure 8I**).

*Cxcl12* was expressed in multiple cell types, including the artery endothelial cells themselves and the right ventricle trabecular myocardium where we observed biased SpA formation (**Figure 5J, K,** and **Supplementary Figure 8F**)^25^. This expression in only one chamber’s trabecular cardiomyocytes was also seen in human hearts, except the chambers were flipped between the species (i.e. human expression was specific to left and mouse to right) (compare **Figures 5J** with **Figure 4K** and **L**).

To test whether SpA placement was influenced by the localized expression in right ventricle trabeculae, we investigated whether SpA branching within the septum was altered in *Cxcl12* heterozygous neonatal mice. In *Cxcl12^DsRed/+^* mice, SpAs were shifted leftward away from the right ventricle trabeculae, and sometimes were only near the left (**Figure 5I** and **L**). These data support a role for increased *Cxcl12* levels in attracting SpAs during development.

Regarding dominance establishment, we frequently observed that one of the original connections, either to the left or right coronary artery, had grown larger than the other side during earlier developmental stages (**Supplementary Figure 8D**). We observed that mature SpAs without immature artery networks emerged at E16.5 and 17.5 in most hearts, suggesting the acquisition of left, right, or co-dominance (**Figure 5H** and **Supplementary Figure 8E** and G)^43^. In fact, when comparing SpA dominance at these stages (E16.5 and 17.5) between *Cxcl12^+/+^*and *Cxcl12^DsRed/+^* embryos, the distributions were similar to those quantified in neonates. Specifically, there was a trend away from right-dominance and towards co-dominance in *Cxcl12^DsRed/+^* embryos (compare **Supplementary Figure 8H** with **Figure 5F**). Together, these data show that *Cxcl12* expression is an attractive force dictating coronary artery placement, and that decreased levels result in altered coronary artery configurations.

## Discussion

Here, we present a combined and stratified multi-ancestry GWAS of coronary artery dominance using genotypes from MVP participants who had undergone angiograms, including large numbers of participants of European, African, or Admixed American descent. State-of-the-art genotyping, imputation, and linear mixed-model analyses of genetic data enabled identification of ten loci with the strongest signal located downstream of *CXCL12.* Genomic analyses identified individual variants with the potential to alter transcriptional regulation. Whole-organ immunofluorescence and spatial transcriptomics showed that dominance is apparent in humans during fetal development when *CXCL12* is expressed in the heart. Evaluation of heterozygous knockout mice demonstrated that *Cxcl12* expression levels influence mouse SpA dominance by imparting an attractive force on artery localization. Taken together, these data support the notion that *CXCL12* expression directly regulates patterning of human coronary dominance during development.

Prior studies in mouse and zebrafish models have demonstrated a role for *Cxcl12* in artery development. Initially discovered as a chemoattractant for hematopoietic cells^44,45^, mouse knockouts deleting *Cxcl12* revealed a requirement for artery development^46^. Mechanistic studies in zebrafish showed that arteries form adjacent to cells expressing *Cxcl12*. In these locations, it acts as a paracrine signal attracting pre-artery ECs that express Cxcl12’s receptor, Cxcr4. Upon activation, pre-artery cells migrate together to assemble into mature arteries or extend artery tips^21^. This same cellular mechanism functions during coronary artery development in mice with *Cxcl12* being expressed in the outflow tract, myocardium, and epicardium^13,22-24^. Mouse studies also demonstrated that *Cxcl12* selectively signals to pre-artery ECs, which are a minor subset of the ECs within the developing capillary plexus^25^. This causes pre-artery ECs within the network to specifically migrate against the direction of blood flow and join to extend arterial vessels, without compromising blood flow^12,47^.

Animal models have also shown that *Cxcl12* stimulates coronary repair and regeneration. Neonatal mouse heart regeneration is enabled after myocardial infarction by capillary induction of *Cxcl12*, which signals to artery cells and guides their migration to a location where they can form natural collateral artery bypasses that support reinnervation^48-50^. In adult mouse hearts, which don’t regenerate, a single localized injection of *CXCL12* stimulates revascularization myocardial recovery^48,51,52^. Similarly, *Cxcl12*-deficient zebrafish fail to fully regenerate their heart following apical resection due to deficient revascularization^22,53^.

With this knowledge, we propose a model where *CXCL12* expression domains in human fetal hearts attract CXCR4+ pre-artery ECs to pattern artery branch growth. This activity could direct RCA development over the posterior left ventricle in most hearts. We also propose that variants associated with left dominance alter transcription factor binding and modestly decrease *CXCL12* transcription. This could stunt or redirect RCA growth so that the LCA eventually extends to compensate (**Figure 6**). In fact, experiments have shown artery compensation to be a prominent feature of the coronary vasculature^54,55^. Future work will aim to experimentally validate all the causal variants and identify the transcription factor activity they effect.

**Figure 6.**
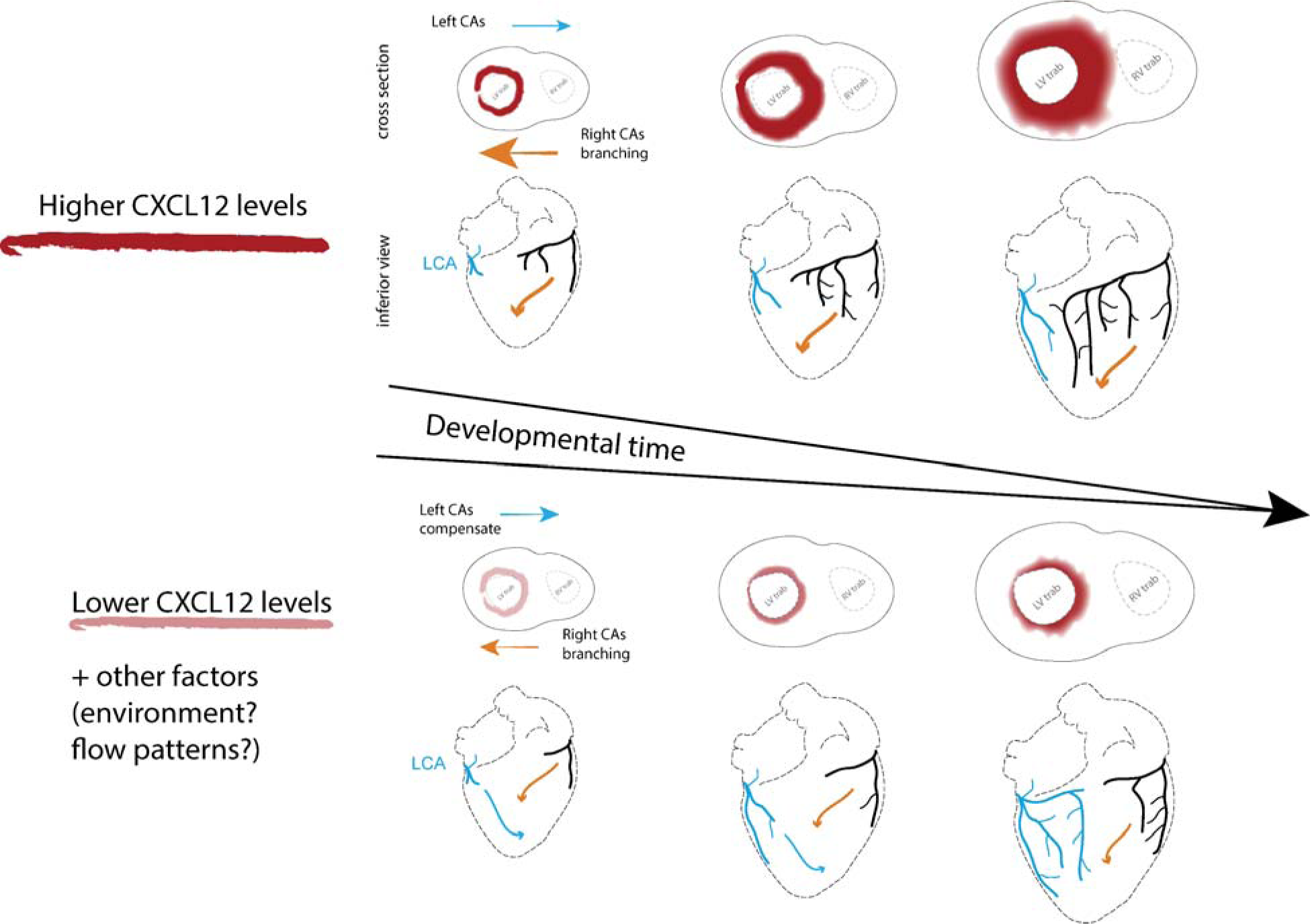
Working model of how *CXCL12* genetic variants influence human coronary artery development and result in different dominance configurations. Genomic data suggested that dominance associated SNPs influence *CXCL12* gene expression with right CA dominance being associated with higher CXCL12 levels. Mouse developmental genetics showed that *Cxcl12* levels influence SpA dominance and indicated that arteries are attracted to form near expression domains. We propose that arterial endothelial cells are attracted to *CXCL12* expression in the epicardium, myocardium, and artery endothelial cells to preferentially extend the right coronary artery on the back of the heart. When *CXCL12* levels are lower, extension of right branches is delayed or misplaced, and the left grows to compensate. Colored arrows indicate the extension path(s) that right (orange) and left (blue) CAs will take based on CXCL12 levels.

Our study stands out among other discovery GWASs in that it identified a susceptibility locus detected in both EUR and AFR populations. Identification of the *CXCL12* locus in both populations, despite EUR imbalance, underscores its importance in determining dominance patterns. A weakness here is the low number of female participants in MVP, although 35% of PMBB participants were female. We will at least in part correct this imbalance when angiogram data becomes available for other biobanks with higher female representation. However, we expect dominance patterning mechanisms to be the same in males and females since dominance ratios are conserved across sexes^56-58^ and genetic data shows an overall genetic correlation of one between sexes for most human traits^59^. We expect to extend discovery in the future when genotyping of additional MVP and PMBB participants is made available, and as additional coronary artery evaluations are performed in participants without angiograms.

A somewhat unanticipated finding from the GWAS was the extensive overlap and colocalization of variants associated with dominance and CAD at the *CXCL12* locus. Whether this finding represents a cascading effect of a single primary action of the gene (vertical pleiotropy) versus an independent effect on multiple traits (horizontal pleiotropy) remains to be determined^60^. In this context, we note that cardiologists have been fascinated by whether dominance impacts heart function or disease since the turn of the 20^th^ century when it was first categorized in postmortem hearts^4^. Many studies have attempted to correlate dominance with cardiovascular events, but no consistent data has emerged^57,58,61-63^. While multiple reports suggest left dominance is associated with a poorer prognosis after a myocardial infarction or revascularization procedures^56-58,64-67^, the clinical relationship between dominance and coronary plaque burden or incident myocardial infarction is less clear^68-70^. To answer these questions more definitively, large scale studies examining the relationship between dominance and an unbiased documentation of atherosclerotic burden need to be conducted, as well as robust and well-powered Mendelian Randomization studies^60,71^.

In summary, our findings provide solid evidence that *CXCL12* patterns human coronary artery growth and branching. Given the ability of *CXCL12* to revascularize the heart through inducing collateral arteries in pre-clinical models, our data is a key piece of evidence increasing confidence that a therapeutic applying localized *CXCL12* could induce collateral arteries in humans. Our work represents an early, but critical, step toward developing a medical revascularization therapy that can one day obviate the need for open heart surgery to provide a coronary artery bypass.

## Materials and Methods

### Population genetic studies

The design of the MVP has been previously described^72^. Briefly, active users of the Veterans Health Administration (VA) of any age have been recruited from more than 60 VA Medical Centers nationwide since 2011 with current enrollment at >1,000,000. Informed consent is obtained from all participants to provide blood for genomic analysis and access to their full EHR within the VA prior to and after enrollment. We linked MVP participants to the Veterans Affairs Clinical Assessment, Reporting, and Tracking (CART) Program, a national quality and safety organization for invasive cardiac procedures, to identify participants who had undergone at least one coronary angiogram by June 2021^73^. Data were available retrospectively starting in 2004 in select sites and from all sites by 2010^74^. The study received ethical and study protocol approval from the VA Central Institutional Review Board.

#### Genetic Data and Quality Control

A total of 658,164 participants enrolled in MVP between 2011 and 2019 were genotyped with a customized Affymetrix Axiom array in three batches. A genotyping quality control (QC) procedure described in detail elsewhere^75^ was applied to all three batches of data and further adapted to remove markers out of Hardy-Weinberg Equilibrium or with differential missingness between sexes. A total of 590,511 autosomal and 12,693 nonPAR X-chromosome markers passing QC were then phased using SHAPEIT4 v4.2.0^76^. Using Minimac4 (v1.0.2)^77^, phased haplotypes were then imputed to a TOPMed panel (GRCh38 reference genome) that included ∼194,000 phased haplotypes.

#### Assignment of coronary dominance

A unique feature of the MVP biobank is its linkage to the Veterans Health Administration (VA) Clinical Assessment, Reporting, and Tracking (CART) Program. This is a national quality and safety initiative that collects the results of all invasive cardiac procedures performed on Veterans within the nationally integrated VA healthcare system, including a structured variable defining coronary dominance. Coronary dominance is collected as a structured variable in the EHR template report used to document the results of coronary angiograms for the CART registry and is typically entered by a member of the team who performed the procedure. One of three dominance options can be selected including right, co-dominant, and left. We first excluded all coronary angiogram reports from individuals with a history of a heart transplant if they were performed on the day of or after the date of their cardiac transplant. For individuals with a single procedure, we assigned dominance based on that single procedure. Most individuals with more than one procedure in CART were fully concordant in their dominance assignments across all procedures. For the remaining, we required that at least 80% of the assignments were concordant to assign a dominance, otherwise the participant was excluded. A total of 61,044 MVP participants were found to have at least one coronary angiogram evaluation with a documentation of dominance.

### Statistical Analysis

#### Genetically inferred ancestry assignment

Population membership to all genotyped participants was assigned using a reference dataset of unrelated individuals from the 1000 Genomes Project (1KGP). This assignment was performed centrally as part of a core MVP project and made available as a core resource for all MVP investigators^78^. Specifically, the smartpca module in the EIGENSOFT package^79^ was used to project the principal components (PCs) loadings from the 1KGP to genotyped MVP participants. To define the genetically similar population, we trained a random forest classifier using cross-population meta-data based on the top 10 PCs from the reference training data. We then used a random forest classifier trained on the predicted MVP PCA data to assign individuals to one of five 1000 Genomes major continental groups including White/Europeans (EUR), non-Hispanic Black/African American (AFR), Admixed American (AMR), East Asians (EAS), and South Asian (SAS). Individuals with random forest probability over 50% for a population were assigned to that population. Those who could not be assigned to a population were assigned to ‘other’. PCs were then generated for all members within each of the five major continental groups.

#### Heritability analyses

We used GREML-LDMS as implemented in Genome-wide Complex Trait Analysis (GCTA) v1.94.1 Linux to estimate the multicomponent narrow sense heritability (h^2^) of dominance. GREML-LDMS is one of the most accurate heritability estimation methods when considering MAF and LD structures that may bias such estimates^80^. Analyses were restricted to GIA groups with at least > 4,000 case subjects with non-right dominance when applying GREML-LDMS to binary traits to ensure an estimate of heritability with an acceptable standard error^81^. Restricted by computing memory requirements, we selected a random sample of right dominance subjects and all non-right dominance subjects at a ratio of 3:1 to run through GREML-LDMS^82,83^. First, SNPs with allele dosage information used for all GWAS were converted to hard-call genotypes using the default settings in PLINK v2.00a3LM. SNPs that were multi-allelic, had MAC < 3, or genotype call-rate < 95% were removed. Since dominance status is a binary trait, SNPs with P-value < 0.05 for Hardy-Weinberg equilibrium or differential missingness in cases (non-right) vs controls (right) were also removed^82,83^. LD scores were calculated separately for each autosome using default GCTA parameters with an r^2^ cutoff of 0.01, and the genome-wide LD score distribution was utilized to allocate SNPs to 1 of 4 LD quartile groups, where groups 1-4 represented SNPs exhibiting increasing LD scores. Within each LD group, SNPs were further stratified into 6 MAF bins ([0.001, 0.01], [0.01, 0.1], [0.1, 0.2], [0.2, 0.3], [0.3, 0.4], [0.4, 0.5]) and a genetic relatedness matrix (GRM) was constructed from each bin, ultimately creating 24 GRMs. Finally, reml function in GCTA was used to fit a model of case status (non-right vs right) based on the 24 GRMs, with age and sex as covariates. Total observed heritability estimates were transformed to estimate disease liability across a range of presumed non-right dominance prevalence estimates in the general population including the observed prevalence in MVP.

#### Genome-wide association study in MVP

We used SAIGE v1.1.6.2^18^ which uses the saddlepoint approximation (SPA) to calibrate unbalanced case-control ratios in score tests based on logistic mixed models and allows for model fitting with either full or sparse genetic relationship matrix (GRM) to conduct the coronary dominance GWASs. Each GWAS was adjusted for sex and 10 principal components where the PCs used for all subjects combined analysis were global and generated using genotype data and PLINK while the PCs used for the analyses stratified by major continental group were generated within each population alone. All summary statistics underwent quality control using EasyQC^84^. Variants were excluded if minor allele count was less than or equal to 6, imputation quality was less than 0.7, or a variant was monomorphic. Sex chromosomes analyses were also excluded.

A locus was considered GWS if at least one lead genetic variant within it reached a *P* < 5×10^-8^ in any of the individual population GWAS or in the combined multi-population GWAS. Since no previous large-scale GWAS of coronary dominance has been reported, all loci reaching GWS were considered novel. We used Functional Mapping and Annotation of Genome-Wide Association Studies (FUMA-GWAS)^76^ to define genomic risk loci including independent, lead, and candidate variants. First, independent genetic variants were identified as variants with a *P* below a specific threshold and not in substantial linkage disequilibrium (LD) with each other (r^2^ < 0.6). Second, variants in LD (r^2^ ≥ 0.6) with an independent variant and with p < 0.05 were retained as candidate variants to form an LD block. Third, LD blocks within 500kb of each other were merged into one locus. Lastly, a second clumping of the independent variants was performed to identify the subset of lead SNPs with LD r^2^ < 0.1 within each locus. For our meta-analyses of Whites alone, we used a UK Biobank release 2b EUR reference panel of genotype data imputed to the UK10K/1000G SNPs created by FUMA including ∼17 million SNPs. This panel includes a random subset of 10,000 unrelated subjects among all subjects with genotype data mapped to the 1000G populations based on the minimum Mahalanobis distance. We used the 1000G AFR reference panel of 661 subjects with ∼43.7 million SNPs for our Black, and the AMR reference panel of 347 subjects with ∼29.5 million SNPs for our Hispanics. We looked up select independently significant variants identified by FUMA in the GTExPortal to further characterize their expression^85^. Lastly, we annotated the lead SNP(s) at each novel locus by creating URL hyperlinks to five variant-base portals: OpenTargets, QTLbase, Common Metabolic Disease Knowledge, Open GWAS, and PhenoScanner.

#### Statistical fine mapping

Statistical fine-mapping was performed using SuSiEx, an accurate and computationally efficient method for cross-population fine-mapping using GWAS summary statistics, which builds on the single-population fine-mapping framework, Sum of Single Effects (SuSiE)^86^. SuSiEx improves the power and resolution of fine-mapping while producing well-calibrated false positive rates and retaining the ability to identify population specific causal variants^20^. We used the 1000 Genomes population phase 3 as reference panel to prioritize SNPs or sets of SNPs (credible set) driving each association signal, accounting for multiple causal variants in a genomic region. We calculated 95% credible sets for each identified signal representing the fewest number of variants whose posterior inclusion probabilities (PIP) for the signal summed to ≥ 0.95. We discarded credible sets in which the variants had a minimum *P* > 1×10^-5^ for the population group in which the signal was mapped.

#### Colocalization

We assessed the presence of colocalization of select genetic association signals for coronary dominance with another relevant trait using COLOC^37^. Evidence of colocalization at a locus with the same causal variant shared between two traits was defined as a posterior probability Bayesian factor H4 (PP.H4.abf) > 0.7 while evidence of colocalization with a different variant was defined as defined as a posterior probability Bayesian factor H3 (PP.H3.abf) > 0.7. We used LocusCompareR to visualize colocalization^87^.

#### Replication in the Penn Medicine Biobank

We used the multi-ancestry Penn Medicine Biobank (PMBB) to replicate select findings in the MVP^88^. Among 43,613 participants with available genomic data in PMBB, 3,141 had coronary angiography procedure data available for analysis. Coronary dominance was ascertained from coronary angiograms performed from 2007-2018 at the Hospital of the University of Pennsylvania (HUP) and Penn Presbyterian Medical Center (PPMC). Angiograms performed at PPMC have coronary dominance documented as a discrete field, which was directly extracted from the electronic health record. Coronary angiograms from HUP have coronary dominance documented as part of a semi-structured free-text report. To determine coronary dominance from HUP angiograms, natural language processing (NLP) was performed using the pre-trained “english-ewt” tree-bank model from the ‘udpip’ package in R^2^. Free-text coronary angiography reports underwent tokenization and lemmatization, and the adjectives (eg. “left”, “right”, “co-dom”) referring to coronary dominance were then extracted using regular expressions.

Assignment of coronary dominance was validated by manual review of 30 randomly sampled coronary angiography reports by a board-certified cardiologist. The NLP algorithm perfectly assigned coronary dominance, with positive- and negative-predictive values of 1. When multiple angiograms were available, we required 80% concordance across studies to assign a coronary dominance phenotype. Although most participants had a high degree of concordance across studies, individuals with <80% concordance across studies were excluded from further analysis. Of the 3,141 subjects, 2730 (87%) were classified by the NLP algorithm as right dominant, 152 (4.8%) as codominant, and 259 (8.2%) as left dominant. Genotyping, imputation, and quality control of PMBB to the TOPMed reference panel has been previously described^88^. GWAS of coronary dominance was performed using the same approach as in MVP adjusted for sex and five genetic principal components.

#### Deep learning models to score and interpret variant effects

##### Model Training

To model cell type-specific chromatin accessibility of human adult cardiac cell types, we trained ChromBPNet models (https://github.com/kundajelab/chrombpnet) on cell-type resolved pseudobulk ATAC-seq profiles. For each of the 87 single-nucleus ATAC-seq datasets in adult human heart from the ENCODE project (https://www.encodeproject.org/search/?searchTerm=snATAC-seq+in+heart+Homo+sapiens+adult+snyder), we subsetted the associated fragments file by cell type based on the ENCODE single cell project’s barcode to cell type mapping. Next, we concatenated the subsetted fragment files across datasets and sorted them to create cell type-specific fragment files for each cell type present in the heart samples. We used the ENCODE peak calling pipeline to call peaks for each cell type based on the corresponding fragments file, and then used these peak and fragment files as inputs for ChromBPNet model training. The peaks and bigwigs used as input to the adult heart cell type ChromBPNet models as well as the ChromBPNet models themselves can be found on Synapse (https://www.synapse.org/Synapse:syn60940384/files/).

To train the ChromBPNet models, we used a 5-fold cross-validation scheme wherein each chromosome was present in the test set of at least one cross-validation fold. The table below shows which chromosomes were a part of the train, validation, and test splits in each fold.

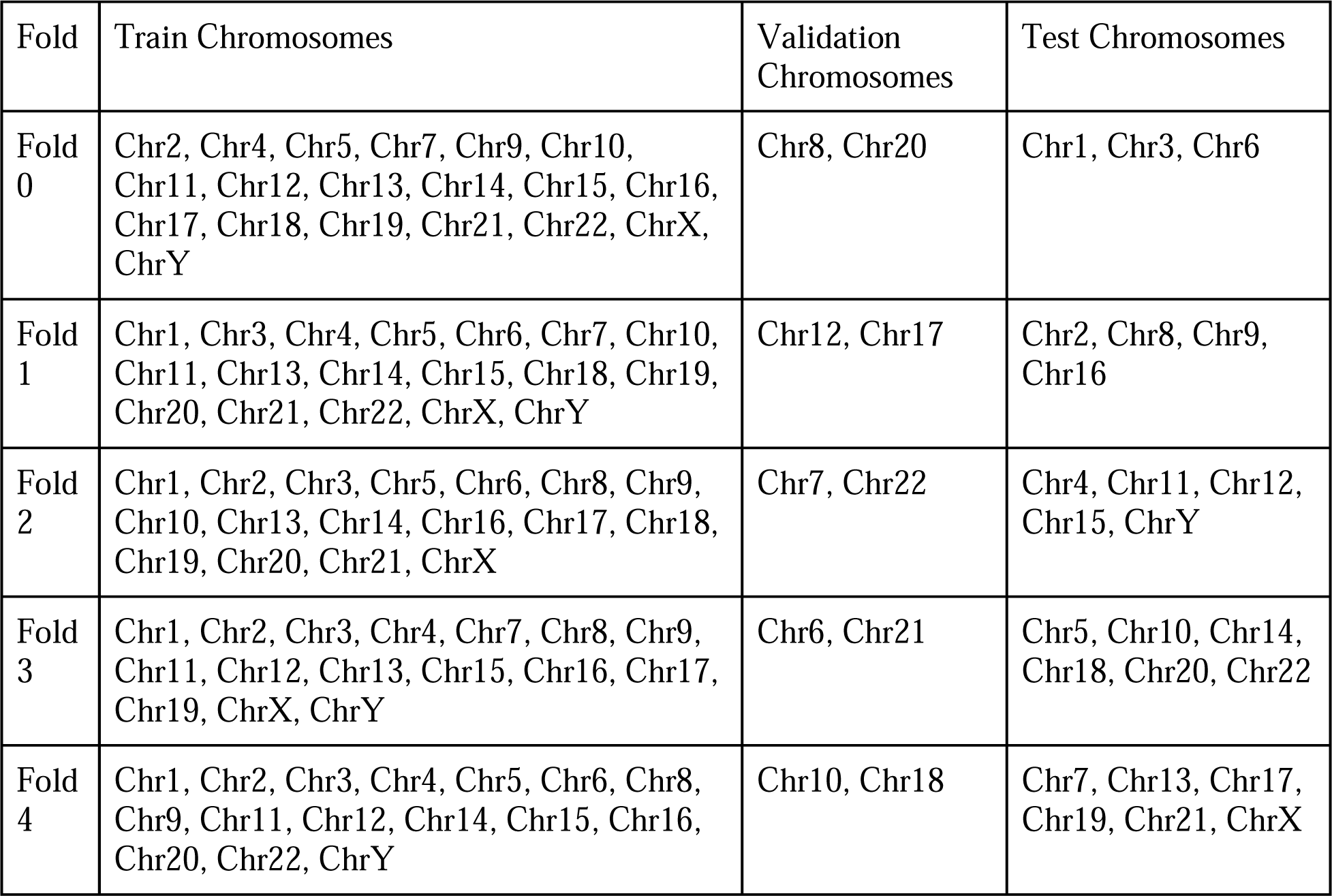

We used the ChromBPNet framework to first train a Tn5 bias model on background regions (not overlapping peaks) from the cardiomyocyte pseudobulk ATAC-seq data. This bias model was used for bias correction while training ChromBPNet models on peaks and background regions from each of the cell types.

To model chromatin accessibility in fetal heart cell types, we trained ChromBPNet models on peak and fragment data from *Ameen et al.* (https://www.sciencedirect.com/science/article/pii/S0092867422015033). The peaks and bigwigs used as input to the fetal heart cell type ChromBPNet models as well as the ChromBPNet models themselves can be found on Synapse (https://www.synapse.org/Synapse:syn60940384/files/). The same 5-fold cross validation scheme was used for the fetal heart models. For Tn5 bias correction on the fetal models, a Tn5 bias model trained on a pseudobulk of ATAC-seq data from all cells (this is represented by the AllCells model). and schemes were used for the fetal heart models, as well.

For each of the fetal and adult heart cell types, we estimated base-resolution predictions of ATAC-seq coverage for each input sequence by averaging predictions from the 5 folds. ChromBPNet predicts two complementary outputs, the total coverage over the input sequence (log (total counts)) and the base-resolution positional probability profiles of reads across the sequence. Hence, we estimate the average total counts as the arithmetic mean of the exponentiated predicted log (total counts) over all 5 folds. We estimate the average of the logits of the predicted profile probabilities per position over the 5 folds, followed by a softmax to obtain the aggregate probability profile. The coverage per position is simply the total coverage x the profile probability at that position.

We computed contribution scores of each base in any input sequence to predicted outputs (log(total counts) or profile probabilities) using the DeepSHAP (https://dl.acm.org/doi/10.5555/3295222.3295230) implementation of the DeepLIFT (https://proceedings.mlr.press/v70/shrikumar17a.html) algorithm, and then averaged scores across all 5 folds of the model, as well.

##### Variant Scoring

To score and identify dominance-associated variants predicted to affect chromatin accessibility, we used the ChromBPNet models for each cell type to predict base-resolution scATAC-seq coverage profiles for 1 kb genomic sequence windows containing reference and alternate alleles of all variants in the dominance GWAS loci. The effect size of each variant was estimated using two measures (1) the log_2_ fold change of the total predicted coverage (total counts) over each sequence window containing the reference and alternate allele (2) the Jensen Shannon distance (JSD) between the base-resolution predicted probability profiles for the ref and alt sequences (this measures a change in the profile shape).

Statistical significance was estimated for both scores on empirical null distributions of variant scores that were computed as follows. We shuffled the 2114 bp sequence centered at each variant multiple times while preserving the dinucleotide frequency, made 2 copies of each shuffled sequence, then inserted each allele of the variant at the center of the shuffled sequence to create a set of 1 million null variants in total. We scored each of these null variants with each model using the same procedure that we used to score each observed variant. Then, for each observed variant, we calculated the proportion of the null variants that had an equally high or higher (more extreme) score to generate an empirical P-value for both the log2 fold change and JSD scores.

This analysis produced a list of dominance-associated variants predicted to cause a statistically significant (absolute log2 fold change or JSD, false discovery rate < 0.01) change in allele-specific read counts. We defined a score called the active-allele-quantile, which is the quantile of the predicted read counts for the allele with the greatest predicted counts at each variant (the active allele), with respect to the distribution of predicted counts for all ATAC-seq peaks in the cell type where the model was trained. Using this score, we further filtered our prioritized variant list to those variants with an active-allele-quantile greater than 0.05, as these variants are likely to either reside in or create an accessible region and therefore are more likely to affect transcription factor binding. The code base for scoring variants is at https://github.com/kundajelab/variant-scorer

### Animal Studies

All mouse colonies were housed and bred in the animal facility at Stanford University in accordance with institutional animal care and use committee (IACUC) guidance on 12 h/12 h day and night cycle with water and food ad libitum.

The following mouse strains were used: C57BL/6J, (Jackson Laboratory, strain code: 000664), CD1 (Charles River, strain 022) and *Cxcl12^DsRed^* (Jackson Laboratory, strain code: 022458). All experiments were conducted in accordance with protocols approved by the Institutional Animal Care and Use (IACUC) Committee of Stanford University.

*Cxcl12^DsRed^* mice were maintained on a C57BL/6J background. To assess SpA dominance, *Cxcl12^DsRed/+^* males were mated with wild type C57BL/6J females to produce litters with wild type (*Cxcl12^+/+^*) and heterozygous (*Cxcl12^DsRed/+^*) mice. All neonatal studies used mice between postnatal days (P)0-P6. For all embryonic studies, timed pregnancies were determined by defining the day on which a vaginal plug was found as E0.5.

#### Assessing SpA dominance

Dominance was assigned from whole-organ images of coronary arteries imaged using light sheet microscopy and obtained by one of two experimental methods, whole-organ immunofluorescence or *in vivo* labeling (see details for each below).

SpA dominance was determined in 3D using Imaris. First, the ventricular septum was identified using the Oblique & Ortho Slicer and Scissors Tools. Then, it was confirmed that the septum was principally irrigated by one or two SpAs. Lastly, using the Oblique & Ortho Slicer and Scissors tools, the SpA was followed until it merged with the most proximal RCA or LCA segments. On rare occasions, the SpA(s) were followed to an independent connection point (ostium) on the aorta. Based on these observations, SpA dominance was scored as right-, left-, or co-dominant, or as aorta. When scoring SpA dominance in *Cxcl12* mice, researcher was blinded to genotype.

#### Whole-organ immunolabeling

Whole heart immunofluorescence was performed following the modified iDISCO+ protocol previously described^39,89^. For all following steps, tissue was always agitated unless noted otherwise. Briefly, animals were perfused with PBS through the dorsal vein, and fixed in 4% paraformaldehyde (PFA)(Electron Microscopy Science 15714) at 4°C for 1hr (embryonic and neonatal hearts) or 2hr (adult hearts), washed 3X in PBS and stored in PBS with 0.01% sodium azide (w/v, Sigma-Aldrich S8032) until ready to process. Hearts were dehydrated in increasing series of methanol/ddH-_2_O dilutions (20%, 40%, 60%, 80%,100% 2X) for 1hr each, followed by overnight incubation in 66% dichloromethane (DCM, Sigma-Aldrich 34856) and 33% methanol. Next, tissue was washed 2X in 100% methanol for 4hrs and bleached overnight at 4°C in 5% hydrogen peroxide (Sigma-Aldrich 216763) in methanol. Next, the hearts are rehydrated in methanol/ddH-_2_O dilutions (80%, 60%, 40%, 20%) for 1hr each, followed by PBS, 0.2% Triton X-100 PBS (2X) and incubated in 20% dimethyl sulfoxide (DMSO), 2.3% Glycine (w/v, Sigma G7126), and 0.2% Triton X-100 PBS at 37°C for 2 days.

For immunofluorescence, hearts were blocked in 10% DMSO, 6% Normal Donkey Serum (NDS, Jackson ImmunoResearch 017-000-121) in 0.2% Triton X-100 for 2 days at 37C. Primary antibodies were prepared in PBS with 5%DMSO, 3% NDS in 0.2% Tween-20 and 0.1% Heparin (w/v, Sigma-Aldrich H3393) and incubated at 37°C for 4-14 days. Antibodies included Cy3 conjugated-αSMA-Cy3 (1:300, Sigma C6198), Podocalyxin (1:1000, R&D Systems MAB1556), RFP (1:1000, Rockland 600-401-379), Jagged1 (Novus, AF599). Secondary antibodies conjugated to either Alexa 555 or 647 (Jackson ImmunoResearch) were matched 1:1 in concentration to their primary target and in prepared in PBS with 3% NDS in 0.2% Tween-20, 0.1% Heparin for the same primary incubation days at 37°C. Washes after each antibody incubation in PBS with 0.2% Tween-20, 0.1% in Heparin were performed in 30min increment until the end of the day, followed by an overnight wash. Before clearing, samples were embedded in 1% low-melting agarose (Sigma-Aldrich A9414) in PBS and dehydrated in methanol/ddH-_2_O dilutions (20%, 40%, 60%, 80%,100% 2X) for 1hr each and 100% overnight. Next, hearts were incubated in 66% DCM and 33% Methanol for 2.5hrs, followed 2X 30min 100% DCM. Finally, samples were cleared in ethyl cinnamate (ECi, Sigma Aldrich 112372), manually inverted a few times, and kept at RT in the dark until imaging.

#### *In vivo* vascular labeling

Neonate pups were gently wrapped in gauze and cooled on ice for 6 minutes to induce hypothermic circulatory arrest. Mice received an intravenous 10 µl retro-orbital injection of Isolectin GS-IB_4_ DyLight™ 649 (1:5, Invitrogen, I32450, in sterile 1X PBS). Neonates were then allowed to recover at 37°C on a warm plate and, when conscious, returned to their mother’s care for 30 min before euthanasia. Hearts were then dissected and fixed in 4% PFA (Electron Microscopy Science, 15714) at 4°C for 1 hour. Subsequently, hearts were washed three times in 1X PBS and stored in cold PBS with 0.01% sodium azide (w/v, Sigma-Aldrich, S8032) and covered from light until ready to process.

Hearts were then cleared following the protocol previously described^90^. For all following steps, tissue was always agitated unless noted otherwise. Before clearing, samples were embedded in 1% low-melting agarose (Sigma-Aldrich, A9414) in PBS and dehydrated in methanol/ddH-_2_O dilutions (20%, 40%, 60%, 80%,100% 2X) for 1hr each and 100% overnight. Next, hearts were incubated in 66% DCM and 33% Methanol overnight, followed next day by 2X 100% DCM washes the next day. Finally, samples were cleared in ethyl cinnamate (ECi, Sigma Aldrich, 112372), manually inverted a few times, and kept at RT in the dark until and after imaging.

#### Light sheet imaging

Samples were imaged with Imspector Pro 7.0.98 software and LaVision BioTec Ultramicroscope II Light sheet microscope in a quartz cuvette filled with ECi. For imaging, we used a MVX10 zoom body (Olympus) with a 2x objective (pixel size of 3.25 µm / x,y) at magnification from 0.63x up to 1.6x. Up to 1400 images were taken for each heart using a z-step size of 3.5µm z step size, and light sheet numerical aperture to 0.111 NA. Band-pass emission filters (mean nm / spread) were used, depending on the excited fluorophores: 525/50 for autofluorescence; 595/40 for Cy3; and 680/30 for AF647. Exposure time was 10ms for single channel and 25ms for multichannel acquisition.

#### Image processing and visualization

Maximum projections were performed using NIH Fiji ImageJ 1.53 software. Representative images in figures were in some cases processed with Despeckle and/or Unsharp Mask features. Raw images are included in Supplement. Imaris 9.5.0 software (Bitplane) was used for 3D rendering. Pixel dimensions were updated from the non-reduced 16-bit image metadata. The Filament Object Tracer module in Imaris was used to generate a simple 3D outline of the main coronary arteries. A root point was manually placed where the RCA and LCA originate from the aorta, i.e. right and left ostium, and filaments were then created and drawn until the farthest length possible using the Autopath and AutoDepth features.

#### Quantitative PCR

Whole hearts were collected from E16.5 mice and lysed in 1 mL of TRIzol (Invitrogen 15596026) in tubes containing Lysing Matrix G (MP Biomedicals: 116916050) with homogenization (3X 30 sec at 30 Hz) in a Qiagen TissueLyser II followed by isolation of the aqueous phase according to the TRIzol reagent user guide. RNA was then extracted and isolated with a NucleoSpin RNA isolation kit (Takara 740955.50) according to the manufacturers’ protocols. 100 ng of total RNA was then reverse transcribed with the SensiFAST cDNA synthesis kit (Meridian Bioscience BIO-65053). Total cDNA was then diluted 1:10 and qPCR was performed with the SensiFAST SYBR® No-ROX Kit (Meridian Bioscience BIO-98005) with 10 μL of reactants in a 384 well plate (ThermoFisher 4483285).

Primer pairs were designed such that they span exon/exon junctions to preclude amplification of genomic DNA and were specific for murine *Cxcl12*. Each well of the plate received 5μL of SensiFAST SYBR® No-ROX Mix, .04 μL of 100mM forward primer, .04 μL of 100mM reverse primer, 2.92μL of water and 2μL of cDNA. After plate preparation with gene specific primers combined in a well with a sample specific cDNA the plates were sealed with MicroAmp™ Optical Adhesive Film (ThermoFisher 4311971) and centrifuged for 5 min at 350g. Plates were run on a Quantstudio Flex 7 (ThermoFisher) with the following parameters: Initial dissociation (2 min at 50°C followed by 10 min at 95°C), 40x PCR stage with SYBR detection (15 sec 95°C, 1 min 60°C) with the final stage being a dissociation step to generate a melt curve for the PCR products to ensure a single product was amplified. During analysis the linear phase of the fluorescence was used to generate Ct (cycle threshold) values. The ddCt (delta-delta Ct) method was used to analyze the results. *Gapdh*, a housekeeping gene, was used as an internal control to normalize between samples which were then compared to the expression of the wild type to quantify *Cxcl12* expression between genotypes. For each well there were 3 technical replicates as well as a water negative control.

#### Embryonic heart analysis

*Cxcl12^DsRed/+^* males were mated with wild type C57BL/6J females to produce litters with wild type (*Cxcl12^+/+^)* and heterozygous (*Cxcl12^DsRed/+^*) embryos. Observance of a copulation plug was considered embryonic day 0.5, and litters were harvested at five embryonic stages (E13.5, 14.5, 15.5, 16.5, and 17.5). At each stage, embryos were fixed in 4% PFA for one hour at 4 degrees and washed in PBS before hearts were dissected. Hearts were next subjected to the whole-organ immunolabeling procedure described above for Jagged1 (arteries) and RFP (Cxcl12 reporter). Whole hearts were then imaged using confocal microscopy and then scored for the SpA anatomy categories reported in **Supplementary Figure 6** (artery network, mature SpA, proximity to ventricular lumen, SpA dominance). For scoring procedures, researchers were blinded to genotype. SpA lengthening in the interventricular septum was scored based on the SpA(s) proximity to the right, left, or both ventricular lumens. In E16.5-17.5 hearts, dominance was determined by tracing mature Jagged1 positive SpA(s) from their origin at the right, left, or both coronary arteries until their terminal end.

#### *Cxcl12* expression analysis

Equal ROIs were placed at the border of each ventricular trabeculae and septum. The area covered by *Cxcl12* reporter signal was measured using Max Entropy threshold option in ImageJ Fiji, then normalized to the total area of ROI.

#### Statistical analysis

Statistical analysis and graphs were generated using Prism 9 (GraphPad). Graphs represent mean values obtained from multiple experiments and error bars represent standard deviation. For qPCR, two-tailed unpaired Student’s t test was used to compare groups within an experiment and the level of significance were assigned to statistics in accordance with their p values (0.05 flagged as ∗, 0.01 flagged as ∗∗, less than 0.001 flagged as ∗∗∗, less than 0.0001 flagged as ∗∗∗∗). When analyzing SpA dominance in *Cxcl12* mice, a two-tailed Chi-square without Yates-correction test was performed in a 2x2 contingency table (right & non-right x wild type & heterozygotes).

### Human Fetal Heart studies

Under IRB approved protocols, human fetal hearts were obtained for developmental analysis^91^. Gestational age was determined by standard dating criteria by last menstrual period and ultrasound^92^. Pregnancies complicated by multiple gestations and known fetal or chromosomal anomalies were excluded. Tissues were processed within 1hr following procedure at which time it was rinsed with cold, sterile PBS. For whole-organ immunolabeling, tissue was fixed in 4% PFA for 4hrs at 4°C before whole-organ immunolabeling. For *in situ* hybridization, hearts were fixed in 4% PFA for 24–48 hr at 4°C, followed by three 15 min washes in PBS.

#### Dominance analysis at fetal stages

3D reconstructions of fetal hearts subjected to whole-organ immunolabeling were performed by the method mentioned above under the heading Image processing and visualization. Briefly, root points were manually placed at the right and left ostia and filaments were then created and drawn until the farthest length possible using the Autopath and AutoDepth features in the Filament Object Tracer module in Imaris. Arterial landmarks and large diameter vessels were prioritized and traced first, followed by vessels extending into the septum on the inferior side of the heart. Right dominance was assigned when the filament trace of the PDA was connected to the RCA filament trace. Additionally, when looking at a central cross-section, the arteries dominating the volume were traced from the RCA and its ostium. Co-dominance was assigned when, looking at a central cross-section, arteries in this volume were equally occupied by those traced from the RCA and LCA to their ostia.

#### *In situ* hybridization

Hearts were sequentially dehydrated in 30%, 50%, 70%, 80%, 90%, and 100% ethanol, washed 3X for 30 min in xylene, washed several times in paraffin, and finally embedded in paraffin which was allowed to harden into a block. For each heart, the whole ventricle was cut into 10-μm-thick sections and captured on glass slides.

*CXCL12* mRNA and α-SMA protein were simultaneously localized on sections using RNAscope® per manufacturers’ instructions. Briefly, slides were baked in 60°C for 1hr in a dry oven. Next, slides were incubated 2X in xylene with slight agitation for 5 min in RT under fume hood. Immediately after, slides were incubated 2X in 100% ethanol for 2 min under the same conditions. Slides were then dried for 5 min in the drying oven at 60°C. Afterwards, ∼5 drops of hydrogen peroxide were added to the deparaffinized slides and incubated at RT for 10 min. Slides were washed 2X with fresh distilled water. In a food steamer, target retrieval was performed for 15 minutes with RNAscope Retrieval Buffer. The tissue was pretreated in mild conditions, with RNAscope Protease III for 15 min. *In situ* hybridization was continued with the RNAscope® Multiplex Fluorescent Reagent Kit v2 (ACD 323100) using the following probes: human *CXCL12* (ACD 422991), positive control (ACD 320861), and negative control (ACD 320871). The ACD RNAscope protocol was followed in Channel 1 for all probes with Opal 650 fluorophore (Akoya FP1496001KT). Prior to mounting, slides were fixed in 4% PFA for 30 min, followed by 3X 15 min washes in PBS-0.1% Tween. The slides were then immunostained with Cy3 conjugated-α-SMA-Cy3 (1:300, Sigma C6198) for 1hr at RT. Then, slides were washed for 10 min 3X in PBS-0.1% Tween. To remove blood cells and reduce autofluorescence, slides were washed in CuSO_4_ for 30 min at RT in a nutating mixer. Slides were then washed with distilled water before adding DAPI and mounting in ProLong Gold Antifade reagent (Invitrogen P36934). Finally, slides were imaged within a week on a confocal microscope (Zeiss LSM 980).

#### Single cell and spatial transcriptomics

Detailed MERFISH and scRNAseq methods are reported elsewhere^41^. Briefly, heart samples were collected at the University of California, San Diego (UCSD) in strict observance of the legal and institutional ethical regulations under IRB-approved protocols.

For single-cell dissociation, tissue samples from eight hearts were enzymatically digested and subjected to FACS before before processing for scRNA-seq. Single-cell droplet libraries were prepared according to the manufacturer’s instructions (10X Genomics) and sequenced using a HiSeq 4000 (Illumina). Raw sequencing reads were processed using the Cell Ranger v3.0.1 pipeline and processed using Seurat v4.0.1. Following standard quality controls, gene expression was normalized for genes expressed per cell and total expression by the NormalizeData function. A nearest neighbor graph was generated and used for graph-based, semi-unsupervised clustering and uniform manifold approximation and projection (UMAP) to project the cells into two dimensions. Cell identities were assigned to the clusters by cross-referencing their marker genes with known cardiac cell type markers from both human and mouse studies.

For MERFISH, the sample was embedded in OCT and sectioned such that all major cardiac structures were captured. MERFISH measurements were taken for 238 genes with 10 non-targeting blank controls using published methods^93,94^. The 238 genes were selected to spatially localize the 75 cell subpopulations present in the above-described scRNAseq dataset. This list was identified using a combination of differentially expressed genes in the 75 cell populations and manual selection of previously characterized cardiac cell type markers^41^.

To integrate the scRNA-seq and MERFISH datasets, the 238 MERFISH genes were utilized with Scanpy’s implementation of Harmony to project both datasets into a shared PCA space before visualization with UMAP. To impute a complete expression profile and cell label for each MERFISH profile, expression profiles and cell labels of the closest scRNA-seq cell were assigned to the MERFISH cells in the Harmony PCA space using the Euclidean distance metric. Performance was evaluated as described^41^.

URLs

GCTA, http://cnsgenomics.com/software/gcta/#Overview;

SAIGE, https://saigegit.github.io/SAIGE-doc/

PLINK: https://www.cog-genomics.org/plink/;

EasyQC, https://www.uni-regensburg.de/medizin/epidemiologie-praeventivmedizin/genetische-epidemiologie/software/;

R statistical software, www.R-project.org;

FUMA, http://fuma.ctglab.nl/;

SuSiEx, https://github.com/getian107/SuSiEx

LocusZoom, https://my.locuszoom.org/

GTExPortal, https://www.gtexportal.org/home/

OpenTargets, https://genetics.opentargets.org/;

QTLbase, http://www.mulinlab.org/qtlbase;

Common Metabolic Disease Knowledge Portal, https://hugeamp.org/;

Open GWAS, https://gwas.mrcieu.ac.uk/;

PhenoScanner, http://www.phenoscanner.medschl.cam.ac.uk/;

CADD, https://cadd.gs.washington.edu/

COLOC, https://chr1swallace.github.io/coloc/articles/a01_intro.html

LDSC, https://github.com/bulik/ldsc;

CARDIoGRAMplusC4D, http://www.cardiogramplusc4d.org;

## Data Availability

The full summary level association statistics from each genetically inferred ancestry population association analyses in MVP from this report will be available through dbGaP, with accession number phs001672 at the time of publication in a peer reviewed journal.

## Acknowledgements

Support for title page creation and format was provided by AuthorArranger, a tool developed at the National Cancer Institute. The Genotype-Tissue Expression (GTEx) Project was supported by the Common Fund of the Office of the Director of the National Institutes of Health, and by NCI, NHGRI, NHLBI, NIDA, NIMH, and NINDS. The data/figures used for the analyses described in this manuscript were obtained from the GTExPortal on 10/18/21.

## Consortia

The VA Million Veteran Program. Members of the consortium can be found in the Supplementary Text File.

## Author Contributions

*Concept and design:* P.E.R.C., D.Z., S.L.C, K.R-H., T.L.A.

*Acquisition, analysis, or interpretation of data:* P.E.R.C., D.Z., J.Z., J.A.N, P.P., A.M.M., E.N.F., X.F., S.K., S.S.D., I.E., P.F.K., A.T.H., S.A, S.P., D.D., S.M.D., K-M.C., M.G.L., V.D.W., A.M.P., M.E.P., S.W.W., P.S.T, A.K., N.C.C., S.L.C, K.R-H., T.L.A.

*Drafting of the manuscript:* P.E.R.C, D.Z., J.Z., S.L.C., K.R-H., T.L.A.

*Critical revision of the manuscript for important intellectual content:* P.E.R.C, D.Z., M.G.L, S.L.C., K.R-H., T.L.A.

## Data and materials availability

The full summary level association statistics from each genetically inferred ancestry population association analyses in MVP from this report will be available through dbGaP, with accession number phs001672 at the time of publication in a peer reviewed journal. All other data are available in the main text or the supplementary materials.

## Supplementary Materials

Figs. S1 to S8

Supplementary Text

Tables S1 to S8

