## Supplemental Figures and Text for "*CXCL12* drives natural variation in coronary artery anatomy across diverse populations"

### *CXCL12* drives natural variation in coronary artery anatomy across diverse populations
SUPPLEMENTARY FIGURES AND TEXT

Pamela E. Rios Coronado[1]†, Daniela Zanetti[2,3,4]†, Jiayan Zhou[3,2]†, Jeffrey A. Naftaly[1], Pratima Prabala[1], Azalia M. Martínez Jaimes[5,1], Elie N. Farah[6], Xiaochen Fan[1], Soumya Kundu[7,8], Salil S. Deshpande[9], Ivy Evergreen[7], Pik Fang Kho[2,3], Austin T. Hilliard[3], Sarah Abramowitz[10,11,12], Saiju Pyarajan[13], Daniel Dochtermann[13], Million Veteran Program[14], Scott M. Damrauer[15,16,17], Kyong-Mi Chang[15,18], Michael G. Levin[10,15], Virginia D. Winn[19], Anca M. Paşca[20], Mary E. Plomondon[21,22], Stephen W. Waldo[21,22,23], Philip S. Tsao[3,24,25], Anshul Kundaje[7,8], Neil C. Chi[6], Shoa L. Clarke[2,3,26]‡, Kristy Red-Horse[1,27,28]*‡, Themistocles L. Assimes[2,3,25,29]*‡

[1]Department of Biology, Stanford University; Stanford, CA, USA
[2]Department of Medicine, Division of Cardiovascular Medicine, Stanford University School of Medicine; Stanford, CA, USA
[3]VA Palo Alto Health Care System; Palo Alto, CA, USA
[4]Institute of Genetic and Biomedical Research, National Research Council; Cagliari, Sardinia, Italy
[5]Department of Developmental Biology, Stanford University School of Medicine; Stanford, CA, USA
[6]Department of Medicine, Division of Cardiology, University of California San Diego; La Jolla, CA, USA
[7]Department of Genetics, Stanford University School of Medicine; Stanford, CA, USA
[8]Department of Computer Science, Stanford University; Stanford, CA, USA
[9]Institute for Computational and Mathematical Engineering, Stanford University School of Medicine; Stanford, CA, USA
[10]Department of Medicine, Division of Cardiovascular Medicine, University of Pennsylvania Perelman School of Medicine; Philadelphia, PA, USA
[11]Sarnoff Cardiovascular Research Foundation; McLean, VA, USA
[12]Donald and Barbara Zucker School of Medicine at Hofstra/Northwell; Hempstead, NY, USA
[13]Center for Data and Computational Sciences, VA Boston Healthcare System; Boston, MA, USA
[14]A list of consortium members can be found in the Supplementary File
[15]Corporal Michael J. Crescenz VA Medical Center; Philadelphia, PA, USA
[16]Department of Surgery, University of Pennsylvania Perelman School of Medicine; Philadelphia, PA, USA
[17]Department of Genetics, University of Pennsylvania Perelman School of Medicine; Philadelphia, PA, USA
[18]Department of Medicine, Division of Gastroenterology and Hepatology, University of Pennsylvania Perelman School of Medicine; Philadelphia, PA, USA
[19]Department of Obstetrics and Gynecology, Stanford University School of Medicine; Stanford, CA, USA
[20]Department of Pediatrics, Neonatology, Stanford University School of Medicine; Stanford, CA, USA
[21]Department of Medicine, Rocky Mountain Regional VA Medical Center; Aurora, CO, USA
[22]CART Program, VHA Office of Quality and Patient Safety; Washington, DC, USA
[23]Division of Cardiology, University of Colorado School of Medicine; Aurora, CO, USA
[24]Department of Medicine, Stanford University School of Medicine; Stanford, CA, USA
[25]Cardiovascular Institute, Stanford University School of Medicine; Stanford, CA, USA
[26]Department of Medicine, Stanford Prevention Research Center, Stanford University School of Medicine; Stanford, CA, USA
[27]Institute for Stem Cell Biology and Regenerative Medicine, Stanford University School of Medicine; Stanford, CA, USA
[28]Howard Hughes Medical Institute; Chevy Chase, MD, USA
[29]Department of Epidemiology and Population Health, Stanford University School of Medicine; Stanford, CA, USA

†Pamela E. Rios Coronado, Daniela Zanetti, and Jiayan Zhou contributed as co-lead junior authors.
‡Shoa L. Clarke, Kristy Red-Horse, and Themistocles L. Assimes contributed as co-supervising senior authors.

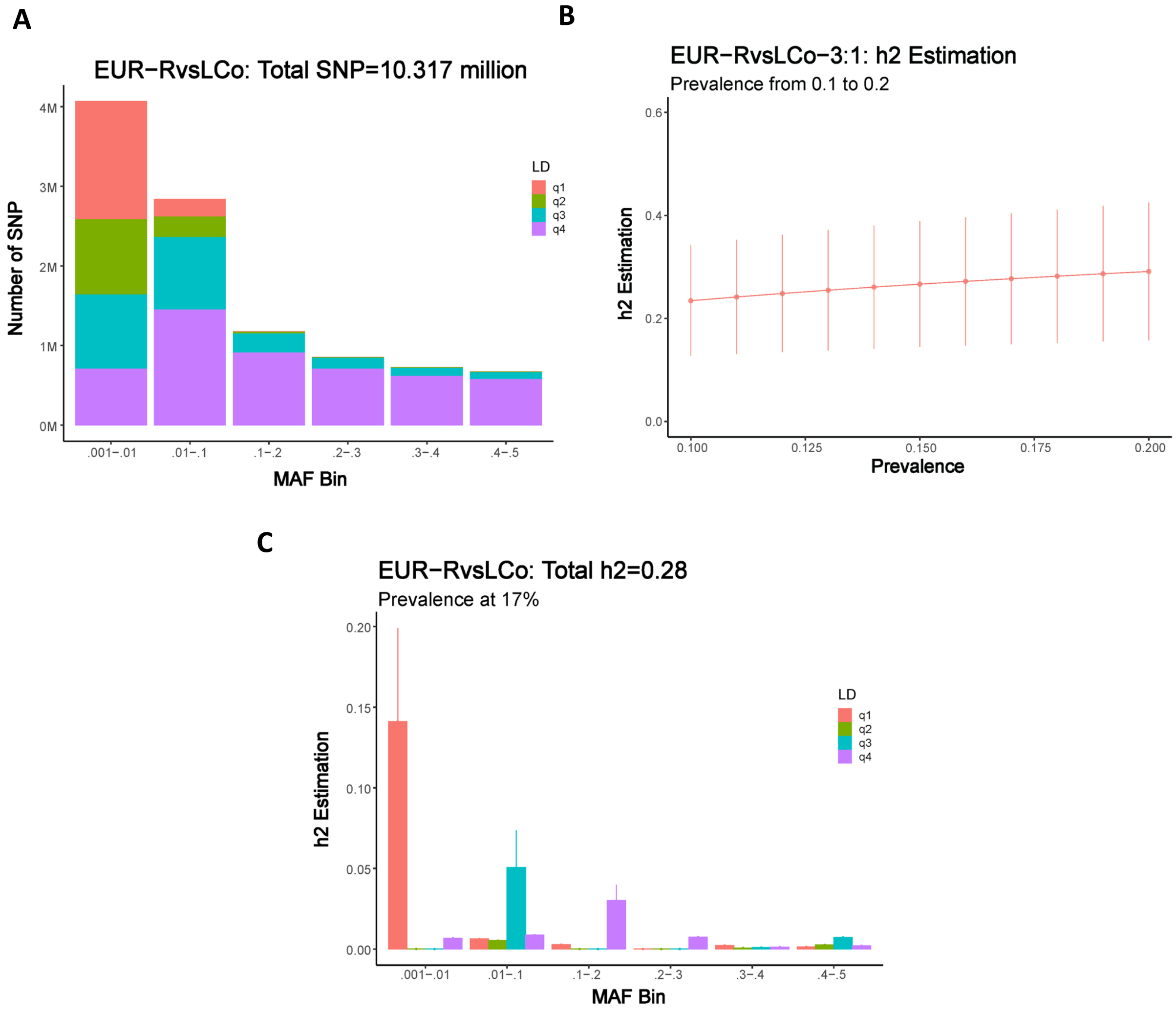


**Supplementary Figure 1. Estimating heritability for coronary artery dominance.** Analyses performed using GREML-LDMS-I as implemented in GCTA for right versus left/co-dominance in European cohort. (**A**) Number of genetic variants by minor allele frequency (MAF) and linkage disequilibrium (LD) score bins (**B**) h2 estimation on a liability scale assuming prevalence of left and co-dominance from 10% to 20% (**C**) Stratified h2 contribution to total h2 by MAD and LD score bins.

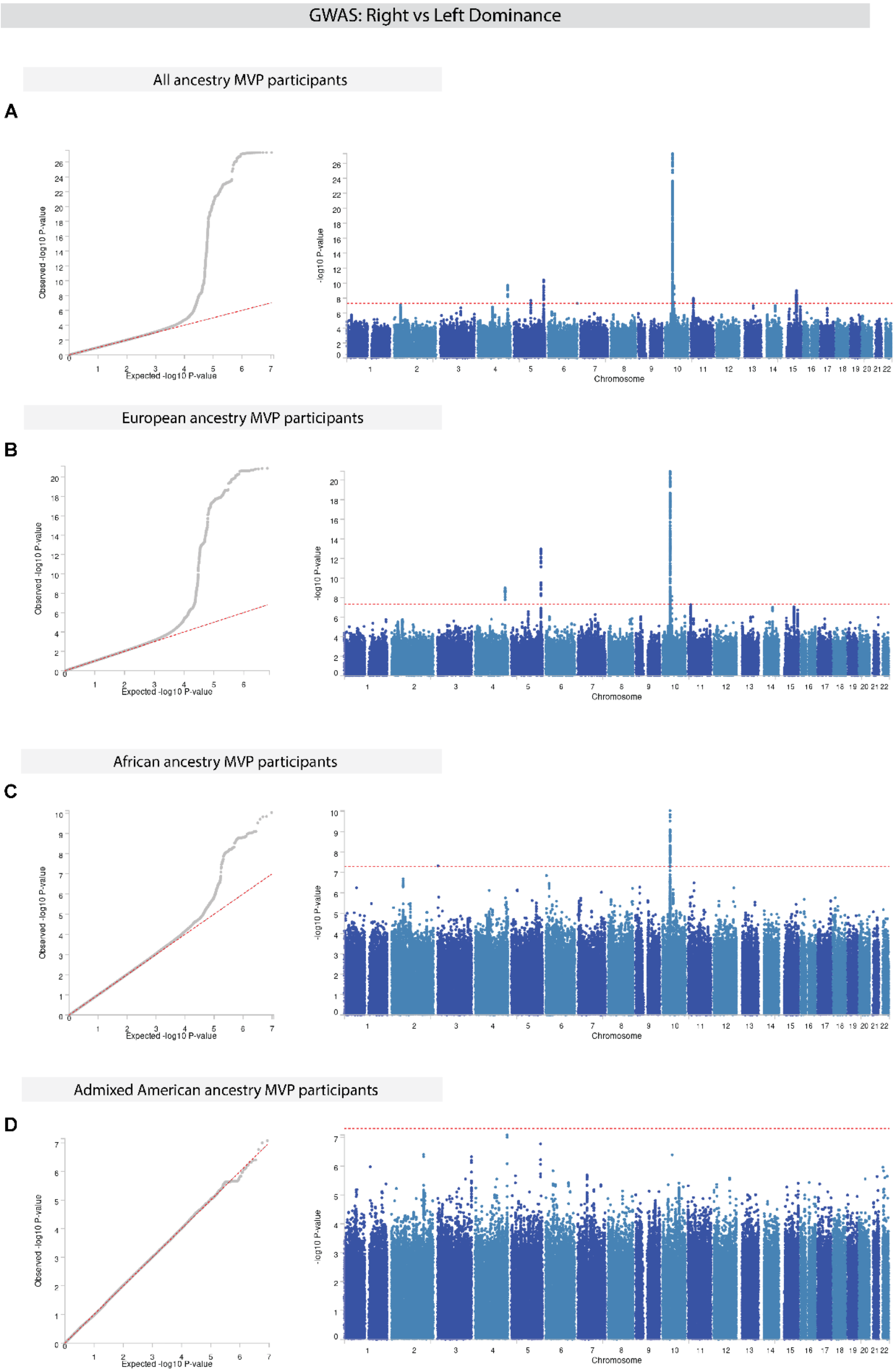


**Supplementary Figure 2**. **Genome-wide association study (GWAS) of right versus left dominance in the Million Veteran Program (MVP).** Q-Q and Manhattan plots for All ancestry (**A**), European (**B**), African (**C**) and Admixed American (**D**) cohorts.

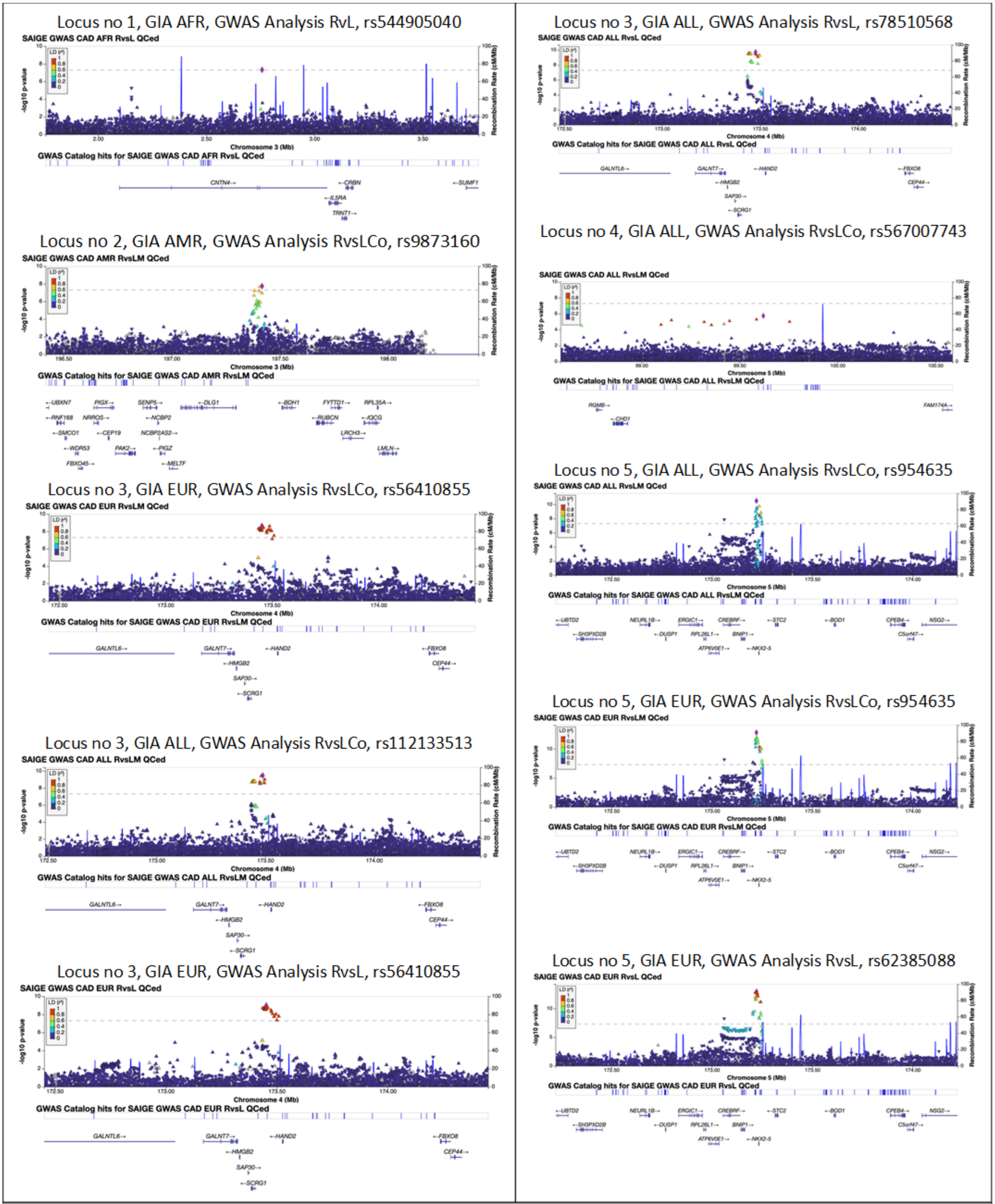


**Supplementary Figure 3. LocusZoom plots around lead genetic variants for analyses listed in Table 1.**

Locus no 5, GIA ALL, GWAS Analysis RvsL, rs5589355

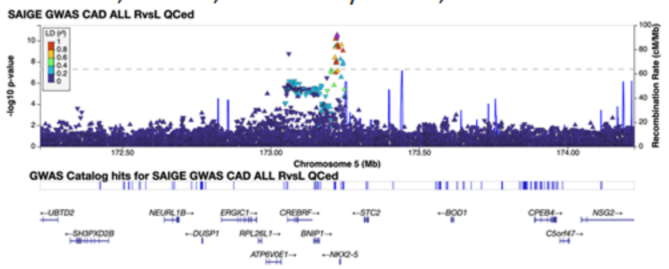


Locus no 6, GIA ALL, GWAS Analysis RvsL, rs2902339

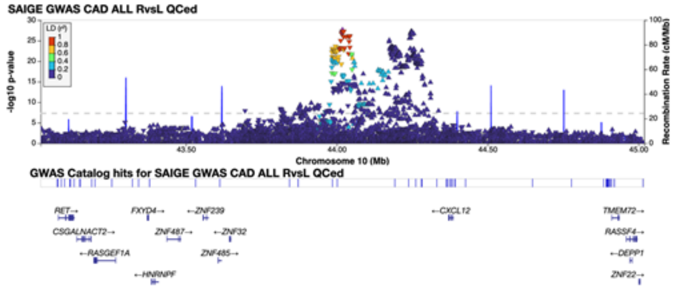


Locus no 6, GIA EUR, GWAS Analysis RvsL, rs7917534

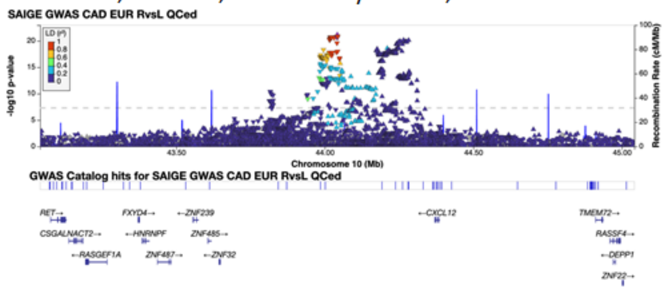


Locus no 6, GIA EUR, GWAS Analysis RvsLCo, rs589655

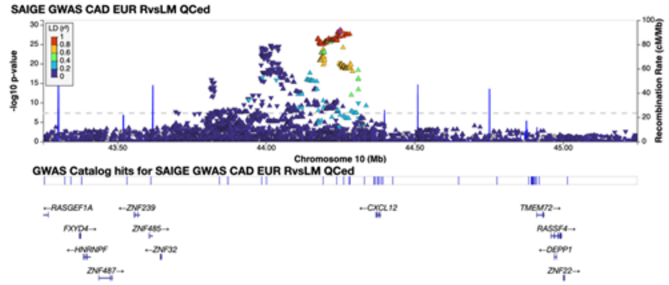


Locus no 6, GIA AFR, GWAS Analysis RvsL, rs2576354

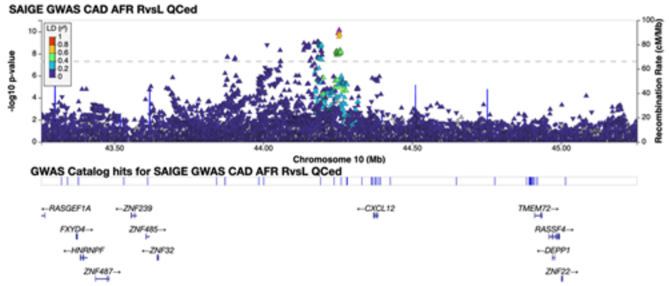


Locus no 6, GIA AFR, GWAS Analysis RvsLCo, rs2576354

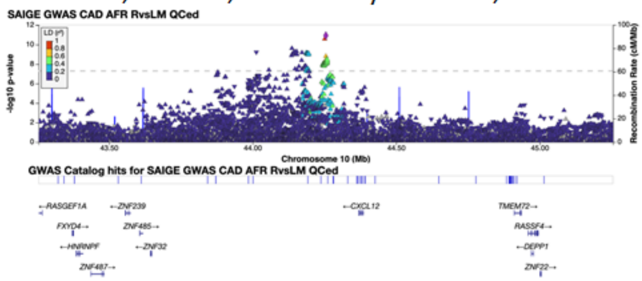


Locus no 6, GIA ALL, GWAS Analysis RvsLCo, rs606314

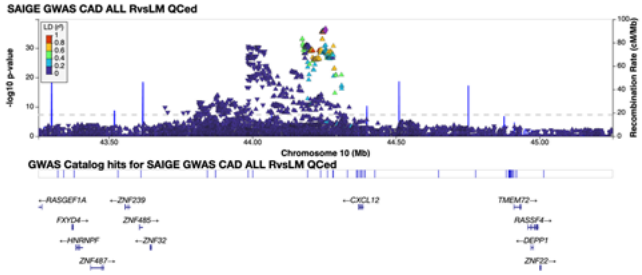


Locus no 6, GIA EUR, GWAS Analysis RvsCo, rs583489

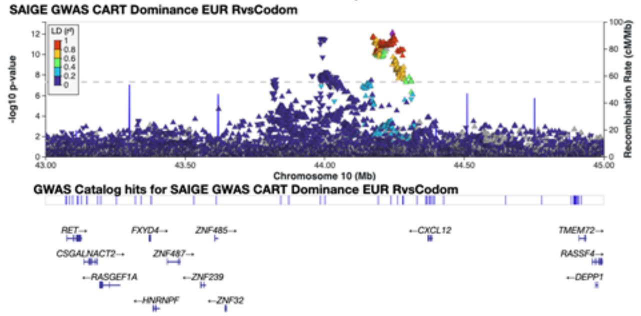


Locus no 6, GIA ALL, GWAS Analysis RvsCo, rs918864065

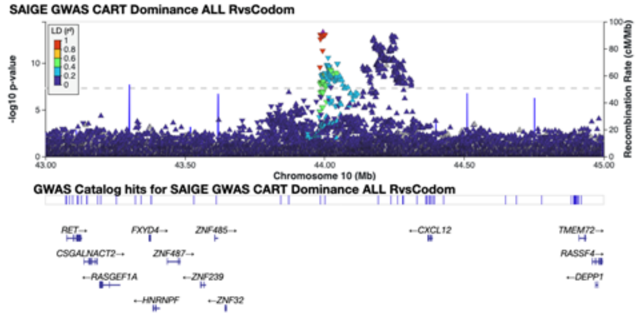


Locus no 7, GIA ALL, GWAS Analysis RvsL, rs1194743

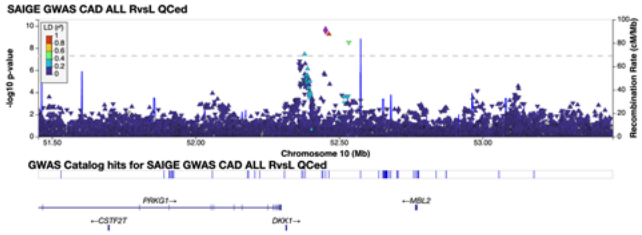


**Supplementary Figure 3 (con't). LocusZoom plots around lead genetic variants for analyses listed in Table 1.**

Locus no 7, GIA EUR, GWAS Analysis RvsL, rs1194743

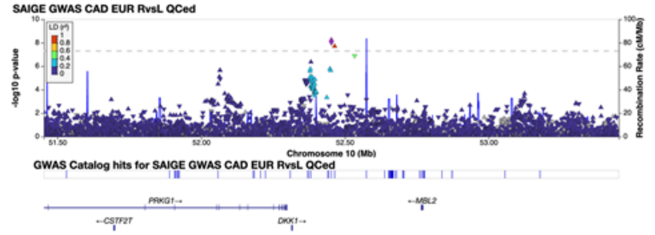


Locus no 8, GIA ALL, GWAS Analysis RvsL, rs35342212

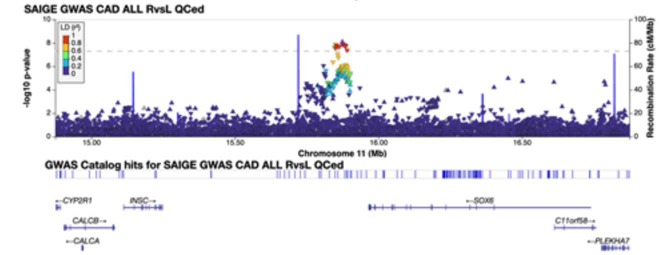


Locus no 8, GIA ALL, GWAS Analysis RvsLCo, rs35342212

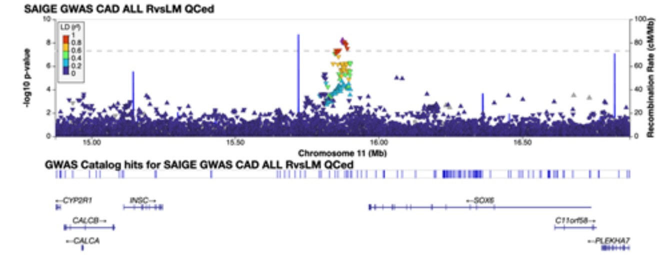


Locus no 9, GIA ALL, GWAS Analysis RvsL, rs58473469

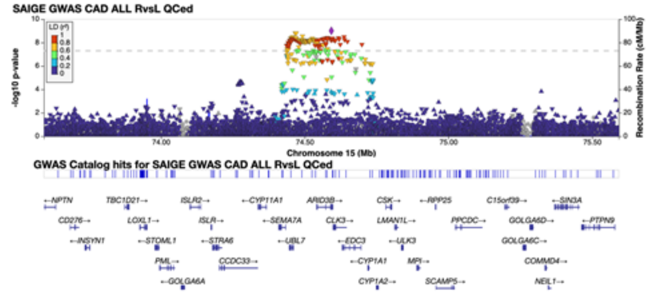


Locus no 10, GIA ALL, GWAS Analysis RvsLCo, rs851058

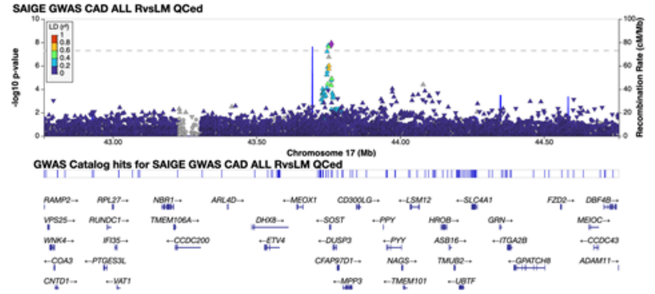


Locus no 10, GIA EUR, GWAS Analysis RvsLCo, rs851058

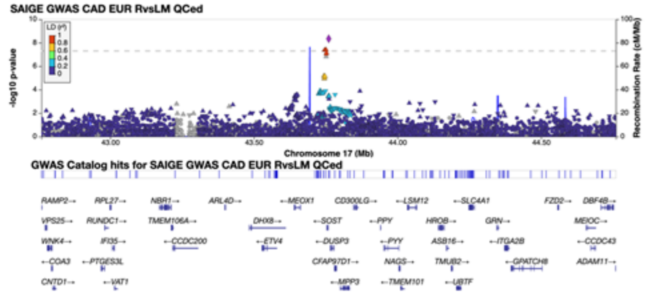


**Supplementary Figure 3 (con't). LocusZoom plots around lead genetic variants for analyses listed in Table 1.**

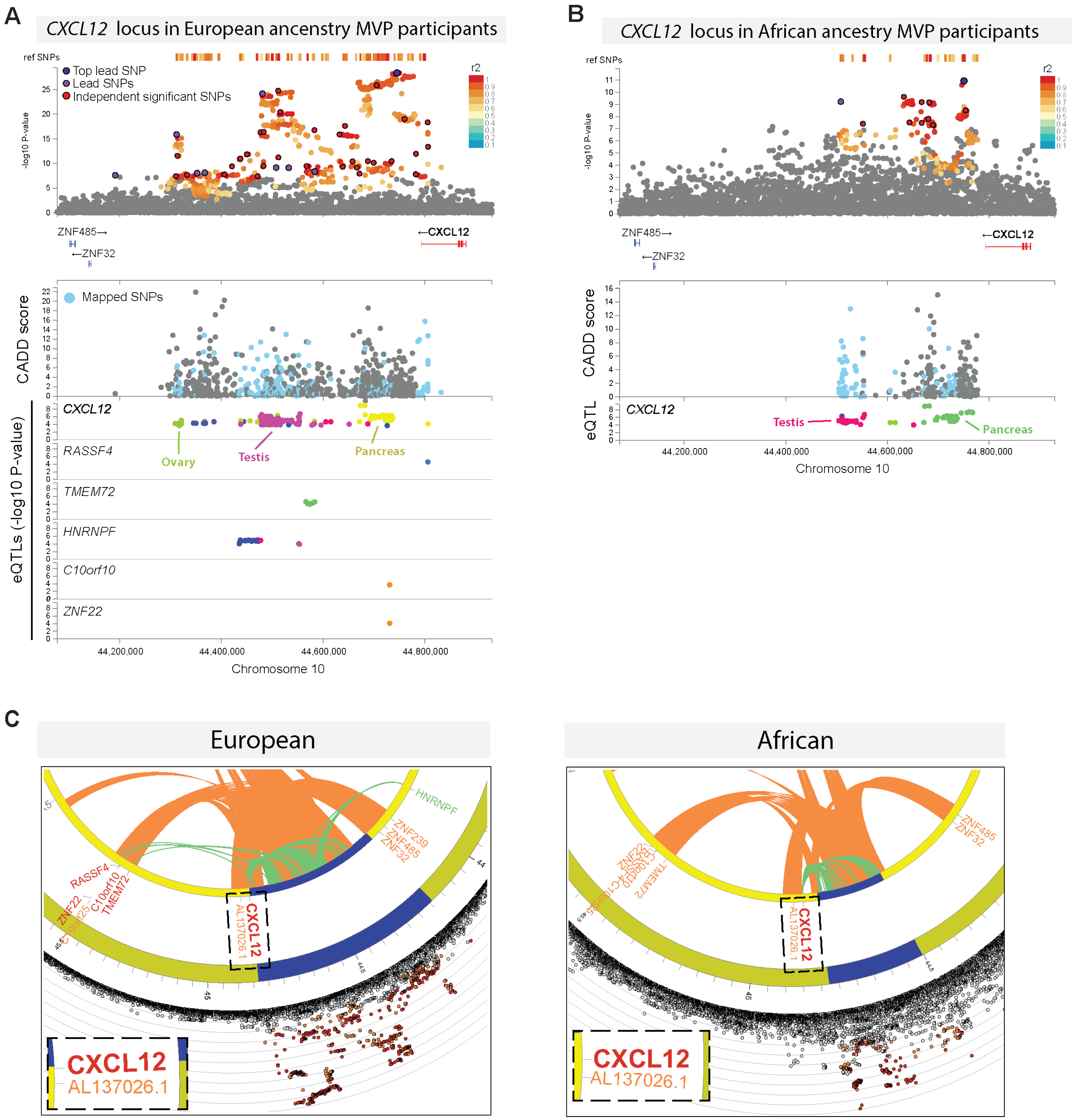


**Supplementary Figure 4. Genomic analyses of *CXCL12* dominance variants. (A and B)** LocusZoom plots derived by FUMA-GWAS in European (**A**) and African (**B**) populations show substantial signal overlap between ancestral groups. SNPs not in LD with any lead or independent significant SNPs are grey. (**A**, **B** bottom panels) CADD scores of SNPs in LD with independent significant SNPs in the locus zoom plots, including SNPs mapped (blue) and not mapped (grey) to genes by eQTLs and/or chromatin interactions. Scores >10 are predicted to affect the expression of nearby genes. Expression quantitative trait loci (eQTL) from the Genotype-Tissue Expression (GTEx) project database are shown for the same variants. eQTLs implicate *CXCL12* and five other genes in European populations, but only *CXCL12* in African populations. (**C**) Circos plots mapping genes by chromatin interactions (orange arcs), eQTLs (green arcs), or both (red gene name). Only *CXCL12* is mapped by both in African populations.

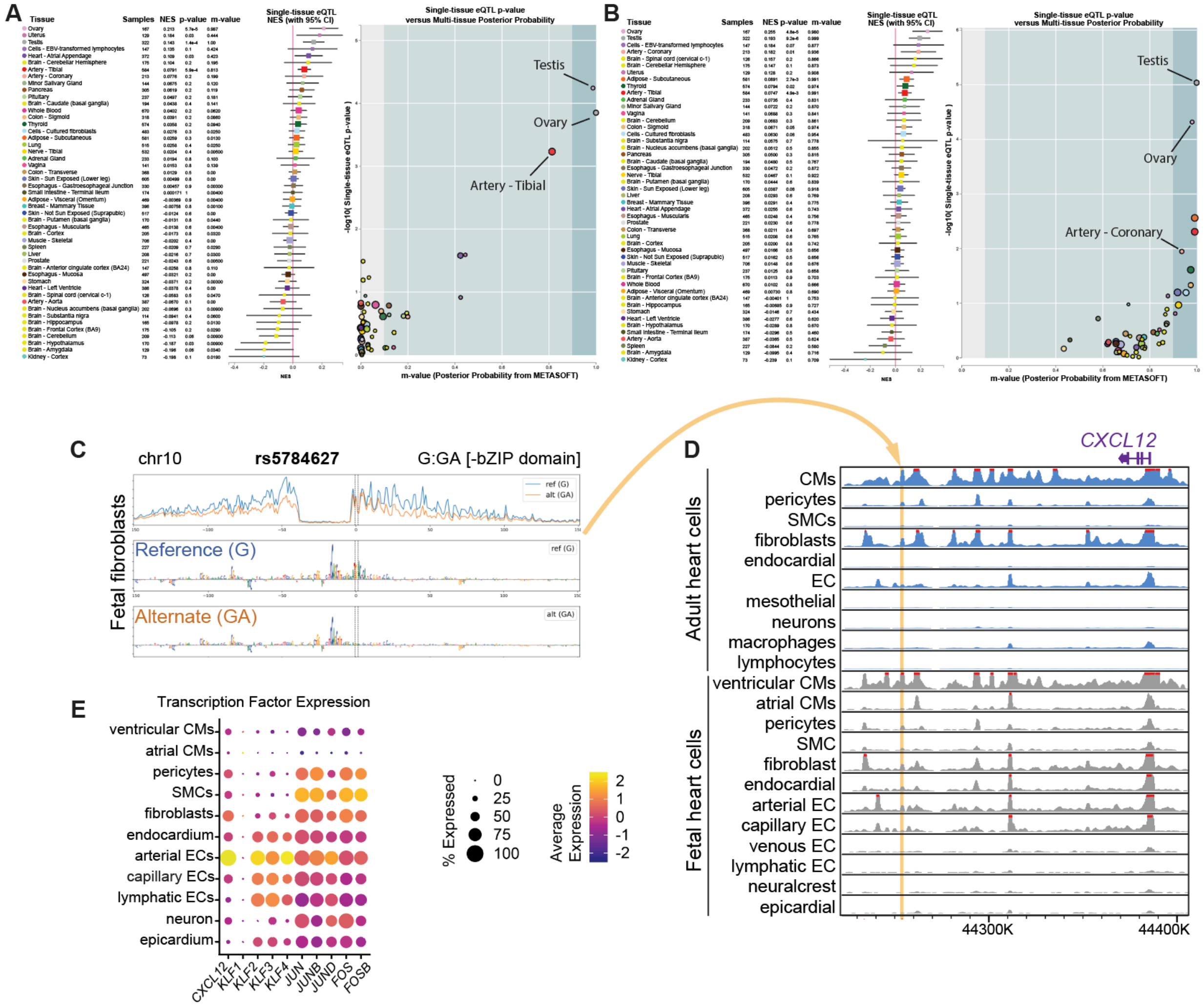


**Supplementary Figure 5. Expression QTLs (eQTLs) and machine learning methods suggest regulatory regions for *CXCL12*. (A** and **B)** eQTL multi-tissue plots from the GTExPortal for two genetic variants, rs1870634 and rs7074248, found to be independently significant for dominance in EUR (**A**) and AFR (**B**) GWAS, respectively. While the strongest effects of tissue expression for these SNPs are in ovary, testis, and the uterus, rs1870634 is also an eQTL for the tibial artery (**A**) and rs7074248 is an eQTL for both tibial and coronary arteries (**B**). (**C**) Predicted per-base accessibility for reference and alternate alleles of rs5784627 suggest a disrupted bZIP domain motif. (**D**) Sequencing tracks of chromatin accessibility near *CXCL12*. Yellow highlights rs5784627 region where open chromatin is coincident with *CXCL12* promoter accessibility in select cardiac cell types, including coronary artery cells. (**E**) scRNAseq from fetal hearts reveals co-expression of *CXCL12* with bZIP domain transcription factors *JUN* and *FOS*.

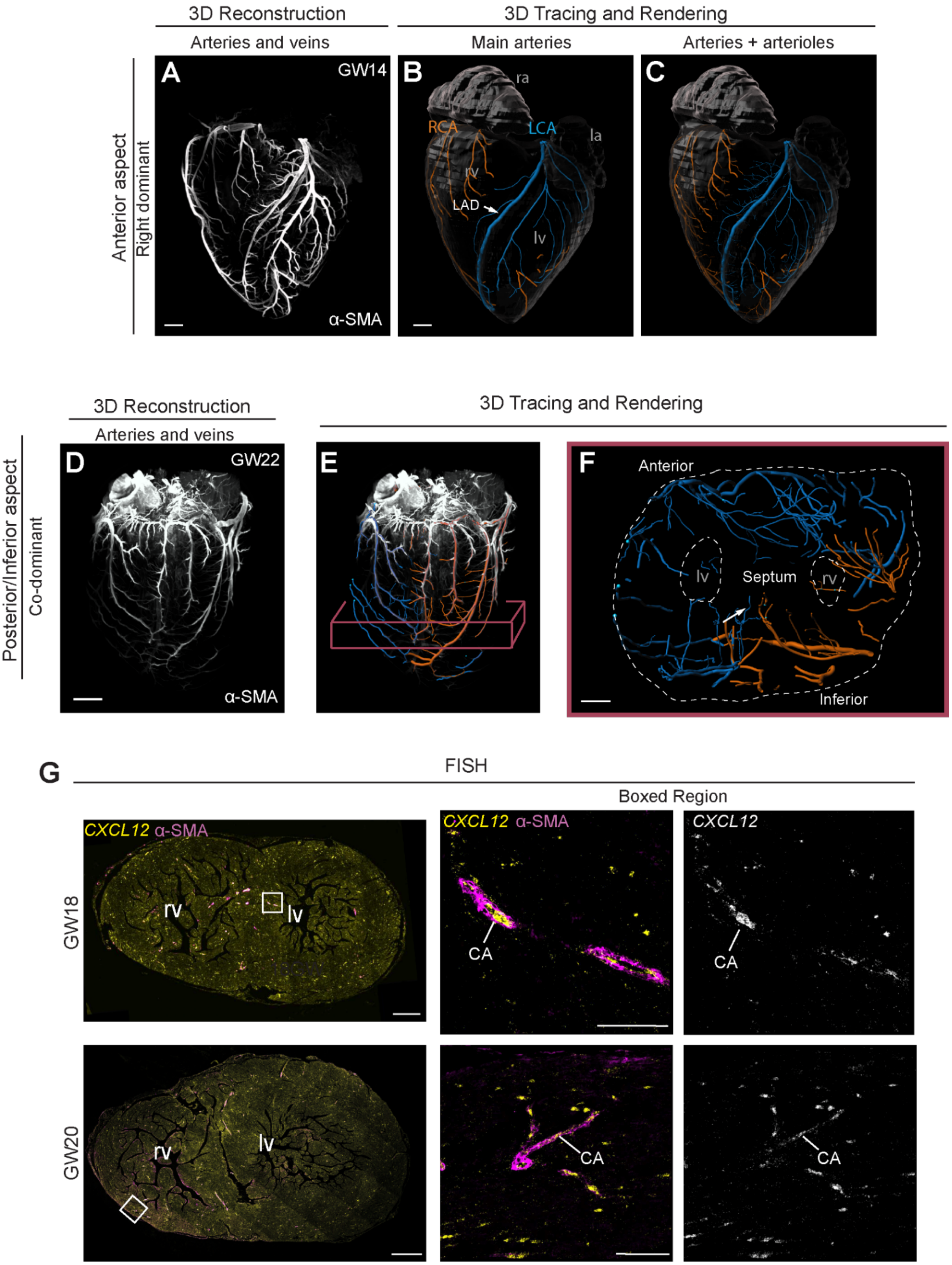
3D Reconstruction
3D Tracing and Rendering
Arteries and veins
Main arteries
Arteries + arterioles
Anterior aspect
Right dominant
A
GW14
α-SMA
B
ra
RCA
LCA
la
rv
LAD
lv
C
3D Reconstruction
3D Tracing and Rendering
Arteries and veins
Posterior/Inferior aspect
Co-dominant
D
GW22
α-SMA
E
F
Anterior
lv
Septum
rv
Inferior
G
FISH
Boxed Region
CXCL12 α-SMA
CXCL12 α-SMA
CXCL12
GW18
rv
lv
CA
CA
GW20
rv
lv
CA
CA

**Supplemental Figure 6. Artery imaging and *CXCL12* expression in fetal hearts.** (**A**) Anterior view of a three-dimensional (3D) reconstruction of a gestational week (GW) 14 heart subjected to whole-organ immunolabeling for alpha-smooth muscle actin (α-SMA). (**B** and **C**) 3D reconstruction of representative tracings of α-SMA+ arteries. Traces highlight branches of the main arteries (**B**) or main with lower order arteries **(C**) that originated from the right (RCA, orange) or left (LCA, blue) coronary ostia where they meet the aorta. (**D-F**) Example of a co-dominant fetal heart as indicated by the presence of both RCA and LCA branches within the IVG. (**E**) and inferior septum (arrow in **F**). (**G**) *CXCL12* fluorescence *in situ* hybridization (FISH) and αSMA immunolabeling on transverse sections through hearts at the indicated ages showed expression in coronary arteries (CA) and other unlabeled cells within the myocardium. LAD, left anterior descending; LCA, left coronary artery; lv, left ventricle; RCA, right coronary artery; rv, right ventricle. Scale bars: A and B 500µm; D, 2mm; F, 1mm; G, left panels, 1mm; middle panels 100µm.

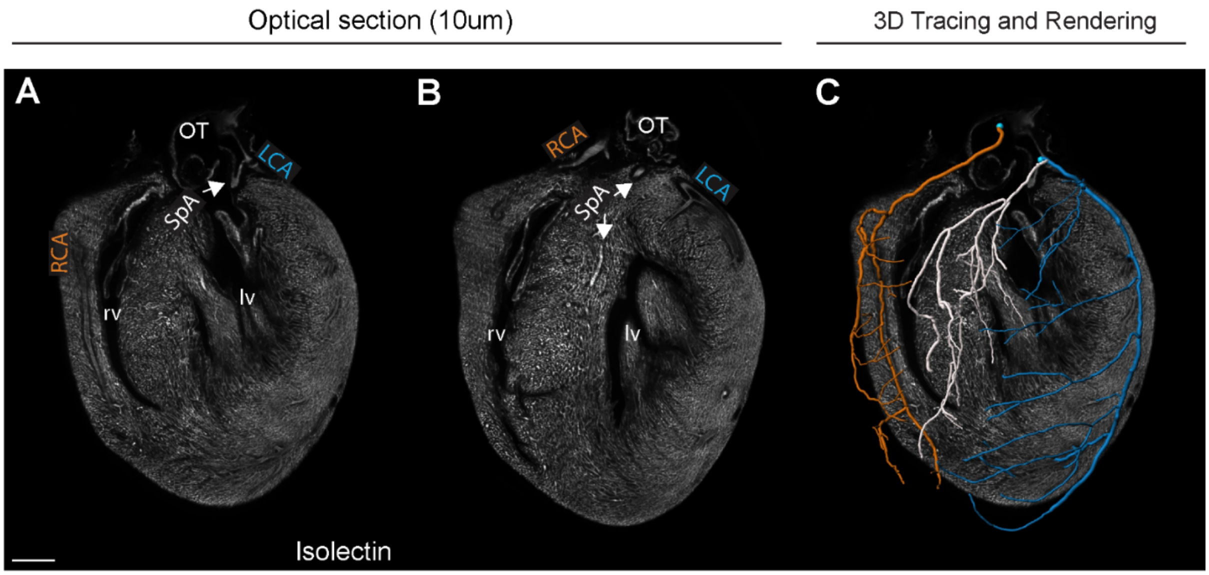


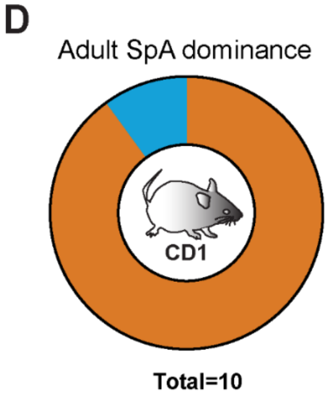


**Supplemental Figure 7. Investigating dominance in mouse hearts.** (**A-C**) Example of the *in vivo* labeling method for identifying septal artery (SpA) dominance in mice. (**D**) Quantification of SpA dominance in adult CD1 mouse hearts revealed that rates are like those observed in neonatal mice. N=10 hearts. LCA, left coronary artery; lv, left ventricle; OT, outflow tract; RCA, right coronary artery; rv, right ventricle. Scale bars: 300µm

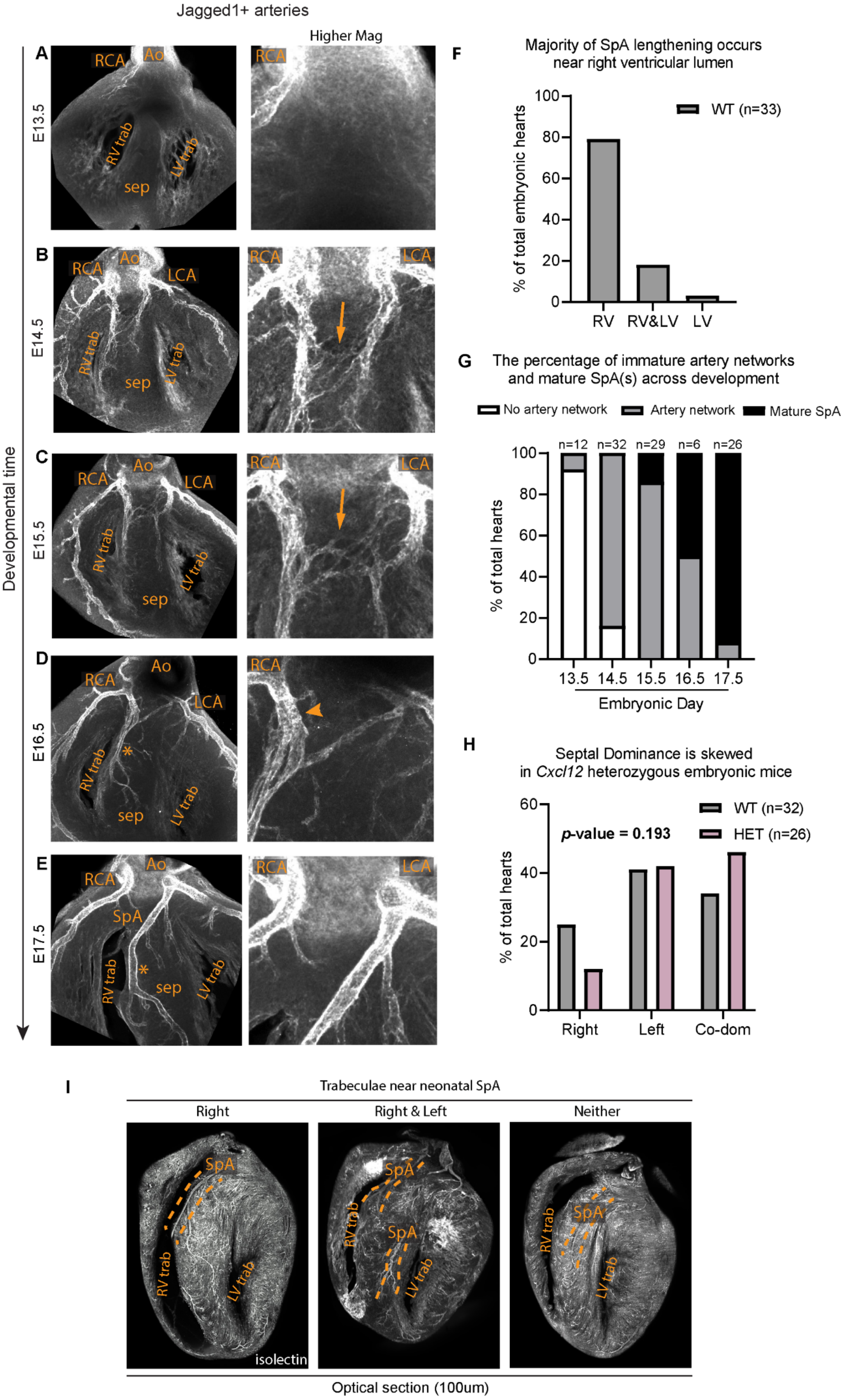

Jagged1+ arteries
Higher Mag
Developmental time
A
E13.5
RCA
Ao
RV trab
LV trab
sep
B
E14.5
RCA
Ao
LCA
RV trab
LV trab
sep
C
E15.5
RCA
Ao
LCA
RV trab
LV trab
sep
D
E16.5
RCA
Ao
LCA
RV trab
LV trab
sep
E
E17.5
RCA
Ao
LCA
SpA
RV trab
LV trab
sep
F
Majority of SpA lengthening occurs near right ventricular lumen
% of total embryonic hearts
WT (n=33)
RV
RV&LV
LV
G
The percentage of immature artery networks and mature SpA(s) across development
No artery network
Artery network
Mature SpA
% of total hearts
n=12
n=32
n=29
n=6
n=26
13.5
14.5
15.5
16.5
17.5
Embryonic Day
H
Septal Dominance is skewed in Cxcl12 heterozygous embryonic mice
p-value = 0.193
WT (n=32)
HET (n=26)
% of total hearts
Right
Left
Co-dom
I
Trabeculae near neonatal SpA
Right
Right & Left
Neither
SpA
RV trab
LV trab
isolectin
Optical section (100um)

**Supplemental Figure 8. Septal artery development in embryonic mouse hearts.** (**A-E**) Whole-organ immunolabeling of wild type mouse hearts, embryonic day (E) 13.5-17.5, with Jagged1 to label artery endothelial cells. Higher magnifications highlight regions where septal arteries (SpA) develop. Arrows indicate interconnected, immature artery precursors in **B** and **C**. Asterisks mark where developing SpAs run along the right ventricular lumen where high *Cxcl12* expression was detected (see **Figure 5I**). Arrowheads point to arteries that have grown larger on one side in **D**, presaging dominance. (**F**) Quantification of SpA growth location with respect to the ventricular lumens show a bias near the right lumen with high *Cxcl12*. (**G**) Quantification of SpA development over developmental time. Dominance in mature SpAs is primarily established between embryonic days 16.5 and 17.5. (**H**) Dominance is skewed in *Cxcl12* heterozygous hearts in embryonic development, similar to postnatal stages (see **Figure 5F**). Data shown for mature SpAs at 16.5 and 17.5. (**I**) Representative images of neonatal SpA localized near right, right and left or neither ventricular trabeculae. Ao, aorta; LCA, left coronary artery; LV trab, left ventricle trabeculae; RCA, right coronary artery; RV trab, right ventricle trabeculae; sep, septum.

**VA Million Veteran Program:**
**Core Acknowledgements for Publications**
**May 2024**

#### MVP Program Office

- Sumitra Muralidhar, Ph.D., Program Director
  US Department of Veterans Affairs, 810 Vermont Avenue NW, Washington, DC 20420
- Jennifer Moser, Ph.D., Associate Director, Scientific Programs
  US Department of Veterans Affairs, 810 Vermont Avenue NW, Washington, DC 20420
- Jennifer E. Deen, B.S., Associate Director, Cohort & Public Relations
  US Department of Veterans Affairs, 810 Vermont Avenue NW, Washington, DC 20420

#### MVP Executive Committee

- Co-Chair: Philip S. Tsao, Ph.D.
  VA Palo Alto Health Care System, 3801 Miranda Avenue, Palo Alto, CA 94304
- Co-Chair: Sumitra Muralidhar, Ph.D.
  US Department of Veterans Affairs, 810 Vermont Avenue NW, Washington, DC 20420
- J. Michael Gaziano, M.D., M.P.H.
  VA Boston Healthcare System, 150 S. Huntington Avenue, Boston, MA 02130
- Elizabeth Hauser, Ph.D.
  Durham VA Medical Center, 508 Fulton Street, Durham, NC 27705
- Amy Kilbourne, Ph.D., M.P.H.
  VA HSR&D, 2215 Fuller Road, Ann Arbor, MI 48105
- Michael Matheny, M.D., M.S., M.P.H.
  VA Tennessee Valley Healthcare System, 1310 24th Ave. South, Nashville, TN 37212
- Dave Oslin, M.D.
  Philadelphia VA Medical Center, 3900 Woodland Avenue, Philadelphia, PA 19104
- Deepak Voora, MD
  Durham VA Medical Center, 508 Fulton Street, Durham, NC 27705

#### MVP Co-Principal Investigators

- J. Michael Gaziano, M.D., M.P.H.
  VA Boston Healthcare System, 150 S. Huntington Avenue, Boston, MA 02130
- Philip S. Tsao, Ph.D.
  VA Palo Alto Health Care System, 3801 Miranda Avenue, Palo Alto, CA 94304

#### MVP Core Operations

- Jessica V. Brewer, M.P.H., Director, MVP Cohort Operations
  VA Boston Healthcare System, 150 S. Huntington Avenue, Boston, MA 02130
- Mary T. Brophy M.D., M.P.H., Director, VA Central Biorepository
  VA Boston Healthcare System, 150 S. Huntington Avenue, Boston, MA 02130
- Kelly Cho, M.P.H, Ph.D., Director, MVP Phenomics
  VA Boston Healthcare System, 150 S. Huntington Avenue, Boston, MA 02130
- Lori Churby, B.S., Director, MVP Regulatory Affairs
  VA Palo Alto Health Care System, 3801 Miranda Avenue, Palo Alto, CA 94304

- Scott L. DuVall, Ph.D., Director, VA Informatics and Computing Infrastructure (VINCI)
  VA Salt Lake City Health Care System, 500 Foothill Drive, Salt Lake City, UT 84148
- Saiju Pyarajan Ph.D., Director, Data and Computational Sciences
  VA Boston Healthcare System, 150 S. Huntington Avenue, Boston, MA 02130
- Robert Ringer, Pharm.D., Director, VA Albuquerque Central Biorepository
  New Mexico VA Health Care System, 1501 San Pedro Drive SE, Albuquerque, NM 87108
- Luis E. Selva, Ph.D., Director, MVP Biorepository Coordination
  VA Boston Healthcare System, 150 S. Huntington Avenue, Boston, MA 02130
- Shahpoor (Alex) Shayan, M.S., Director, MVP PRE Informatics
  VA Boston Healthcare System, 150 S. Huntington Avenue, Boston, MA 02130
- Brady Stephens, M.S., Principal Investigator, MVP Information Center
  Canandaigua VA Medical Center, 400 Fort Hill Avenue, Canandaigua, NY 14424
- Stacey B. Whitbourne, Ph.D., Director, MVP Cohort Development and Management
  VA Boston Healthcare System, 150 S. Huntington Avenue, Boston, MA 02130

**MVP Publications and Presentations Committee**

- Co-Chair: Themistocles L. Assimes, M.D., Ph. D
  VA Palo Alto Health Care System, 3801 Miranda Avenue, Palo Alto, CA 94304
- Co-Chair: Adriana Hung, M.D.; M.P.H
  VA Tennessee Valley Healthcare System, 1310 $24^{th}$ Ave. South, Nashville, TN 37212
- Co-Chair: Henry Kranzler, M.D.
  Philadelphia VA Medical Center, 3900 Woodland Avenue, Philadelphia, PA 19104
